# Excess mortality after mental disorder diagnosis within and outside specialist psychiatric care: nationwide cohort study

**DOI:** 10.64898/2026.09.02.26362001

**Authors:** Kimmo Suokas, Mai Gutvilig, Kaisla Komulainen, Jussi Alho, John J. McGrath, Sami Pirkola, Sonja Lumme, Marko Elovainio, Christian Hakulinen

**Affiliations:** University of Helsinki, Department of Psychology, Faculty of Medicine, Helsinki, Finland. Address: University of Helsinki, Department of Psychology, Faculty of Medicine, PL 21 (Haartmaninkatu 3), FI-00014 Helsingin yliopisto, Finland; Finnish Institute for Health and Welfare, Helsinki, Finland. Address: Finnish Institute for Health and Welfare, P.O. Box 30, FI-00271 Helsinki, Finland; Queensland Centre for Mental Health Research, Brisbane, QLD, Australia. Address: Queensland Centre for Mental Health Research, The Park Centre for Mental Health, Wacol, QLD 4076, Australia; Queensland Brain Institute, University of Queensland, Brisbane, QLD, Australia. Address: Queensland Brain Institute, University of Queensland, Brisbane, QLD 4072, Australia; Tampere University, Faculty of Social Sciences, Tampere, Finland. Address: Tampere University, Faculty of Social Sciences, Arvo Ylpön katu 34 (Arvo 1), FI-33014 Tampere University, Finland; The Pirkanmaa Wellbeing Services County, Department of Psychiatry, Tampere, Finland

**Author notes:** Address: The Pirkanmaa Wellbeing Services County, Department of Psychiatry, P.O. Box 272, FI-33101 Tampere, Finland. **Corresponding:** Kimmo Suokas, MD, PhD, University of Helsinki, PO Box 21, 00014 Helsinki, Finland.

**Keywords:** Primary care, Schizophrenia, Bipolar disorder, Depression, Anxiety

## Abstract

**Background:** People with mental disorders have shortened life expectancy, but much evidence on excess mortality comes from specialist psychiatric populations. We examined variation across the broader diagnosed population by clinical context and time since diagnosis.

**Methods:** We followed 5,526,599 Finnish residents aged 5–95 years without recorded mental disorders at entry from 2011 to 2023 (63 million person-years). ICD-11-grouped diagnoses and mortality rate ratios (MRRs) were assessed by time since diagnosis, psychiatric care setting, substance use disorder (SUD) status, age, and cause of death.

**Results:** During follow-up, 1,463,064 individuals (26.5%) received a diagnosis; the overall adjusted MRR was 2.65 (95% CI 2.53–2.77). However, among diagnosed individuals aged 5–64 years, 46.1% belonged to a subgroup with rapidly declining excess mortality and little evidence of excess beyond two years. This subgroup had depressive, anxiety or fear-related, or stress-related disorders without diagnosed SUD or specialist psychiatric care during follow-up. At two years, MRRs ranged from 0.88 (95% CI 0.73–1.06) to 1.04 (0.88–1.22) in men and 0.92 (0.79–1.07) to 1.12 (0.96–1.31) in women. Excess mortality persisted in specialist care and was particularly high with SUD. It also persisted without SUD or specialist care in schizophrenia and other primary psychotic disorders and older adults with depression.

**Conclusions:** Excess mortality after mental disorder diagnosis may be transient in large parts of the diagnosed population while remaining persistent in others with the same diagnosis. Recognizing these differences is important for interpreting mortality estimates and communicating prognosis across health care.

## Introduction

People with mental disorders are commonly reported to live 7–15 years less than the general population, making premature mortality a major public health concern in psychiatry [1–3]. Population-based evidence, however, suggests that the lifetime probability of receiving a mental disorder diagnosis may approach 70–80% [4–6], while much of the evidence on excess mortality has been derived from specialist psychiatric populations [1–3,7]. What, then, does a single mortality gap tell us about prognosis across the broad population with mental disorders?

Depressive, anxiety, and stress-related disorders are among the most common mental disorders, yet estimates of their associated mortality vary markedly across studies. Recent meta-analyses report increased mortality across these disorders and particularly high mortality shortly after depression diagnosis, but also substantial unexplained heterogeneity in risk estimates [8,9]. Two potentially important dimensions of this clinical context are clinical setting and comorbid substance use disorder (SUD) [10,11]. Both identify populations with substantially different clinical characteristics and mortality risk, but these differences are obscured by average diagnosis-specific estimates. Establishing whether excess mortality persists across the broader diagnosed population or is concentrated in particular clinical groups and periods after diagnosis is important for understanding the mortality gap and for informing patients and clinicians about how mortality risk changes after diagnosis.

We therefore used nationwide data covering both primary and specialist care to examine excess mortality across ICD-11 mental, behavioural, and neurodevelopmental disorder groups according to age, psychiatric care setting, comorbid SUD, cause of death, and time since diagnosis. We further estimated absolute survival loss over ten years. Our aim was to determine whether excess mortality persists broadly across the diagnosed population or varies according to clinical context and time since diagnosis.

## Methods

### Study design and population

This nationwide register-based cohort study included all individuals aged 5–95 years residing in Finland between January 1, 2011, and December 31, 2023. Individuals with prevalent mental disorders before cohort entry were excluded. Participants were followed from cohort entry until death, permanent emigration, or end of follow-up on Dec 31, 2023, whichever occurred first.

Statistics Finland population data were linked using unique personal identity codes to the Care Register for Health Care and Register of Primary Health Care Visits maintained by the Finnish Institute for Health and Welfare (THL). These registers include psychiatric inpatient care since 1975, specialist outpatient care since 1998, and public primary care since 2011 and have shown good coverage and diagnostic validity [12,13].

The study received favorable ethical review from the THL Institutional Review Board (decision #10/2016§751), and data linkage was authorized by Statistics Finland (TK–53–1696-16) and THL. Informed consent is not required for register-based studies in Finland.

### Measures

Diagnoses recorded using ICD-8, ICD-9, or the International Classification of Primary Care, Second Edition were converted to corresponding ICD-10 categories, and register data were preprocessed to maximize diagnostic consistency across data sources and classification systems [13]. Mental, behavioural, and neurodevelopmental disorders were then classified according to ICD-11 groupings using WHO ICD-10-to-ICD-11 mapping tables. Mutually exclusive groupings were created using 10to11MapToOneCategory mappings (see Supplementary Methods).

Mental disorder diagnoses were treated as time-varying. For each diagnostic group, individuals contributed person-time without the diagnosis until the first recorded diagnosis and with the diagnosis thereafter. Following diagnosis, person-time was dynamically allocated to 4 mutually exclusive states according to psychiatric care setting and substance use disorder (SUD) status: outside specialist psychiatric care without SUD, outside specialist psychiatric care with SUD, specialist psychiatric care without SUD, and specialist psychiatric care with SUD. Individuals entered the specialist-care state when the diagnosis was first recorded in specialist psychiatric care and the SUD state at first recorded SUD diagnosis; both classifications were cumulative. Care outside specialist psychiatric care included primary care and nonpsychiatric specialist services. SUD was defined as ICD-11 disorders due to substance use (6C4).

Follow-up was split according to time-varying covariates using a Lexis approach. Age was split into 5-year bands (1-year bands for age-specific analyses), time since diagnosis at 0.25, 0.5, 1, and 2 years and at 2-year intervals thereafter through 10 years, and calendar time at January 1, 2015, and January 1, 2020.

### Covariates

Gender was obtained from the population register and classified as man or woman according to legal gender recorded in administrative registers. Region was classified into 5 Finnish collaborative areas for health care and social welfare services.

Physical comorbidity was measured using the Charlson Comorbidity Index (CCI), a weighted index of chronic medical conditions [14]. Scores were calculated from ICD-10 diagnoses recorded in specialist care during the 5 years before cohort entry and categorized as 0, 1 to 3, or 4 or greater [15]. Age was excluded from the CCI because it was adjusted for separately.

### Outcomes

The primary outcome was all-cause mortality, obtained from Statistics Finland’s Cause of Death Register. Cause-specific deaths were classified as natural or external according to the national classification of causes of death. External deaths included suicides, assaults, and accidents; deaths with missing cause-of-death information were classified separately.

### Statistical analysis

Person-time was aggregated across exposure states, covariates, and time scales. Mortality rates and mortality rate ratios (MRRs) with 95% CIs were estimated using Poisson regression with log person-years as the offset and robust standard errors. The basic model adjusted for age band, calendar period, region, and gender; the second model additionally adjusted for CCI category. MRRs were also estimated according to psychiatric care setting and SUD status, separately for women and men.

We estimated MRRs for all mental, behavioural, and neurodevelopmental disorders combined and by ICD-11 diagnostic group. Diagnosis-specific analyses compared individuals with each diagnosis with all individuals without that specific diagnosis, including those with other mental disorder diagnoses.

Age-specific MRRs were estimated using natural cubic splines for age (4 degrees of freedom, selected by visual inspection of model fit). We compared estimates based on specialist psychiatric diagnoses with those including primary and specialist care to examine the influence of case ascertainment across age.

Analyses according to psychiatric care setting and SUD status were conducted separately for ages 5–64 and 65–95 years because of differences in underlying mortality and disorder distribution; analyses for older individuals are reported in the Supplement. Individuals with SUD before the diagnosis of interest were classified in the corresponding SUD state at diagnosis.

Changes in excess mortality after diagnosis were modeled using interactions between exposure state and natural cubic splines for time since diagnosis (3 degrees of freedom), with model-predicted MRRs evaluated on a 0.1-year grid.

Ten-year restricted mean survival time (RMST) was estimated from CCI-adjusted Poisson models including interactions between exposure state and time since diagnosis. Differences in RMST between individuals with and without each diagnostic group were used to quantify absolute differences in predicted survival during the first ten years after diagnosis. CIs were estimated using nonparametric bootstrap resampling with 200 iterations. Cause-specific RMST differences were estimated separately for natural and external deaths. Additional analyses restricted incident disorders to 2016 onward to reduce potential prevalent-case misclassification and repeated analyses using ICD-10 main diagnostic groups. Analyses were conducted from February through August 2026 using R version 4.4.3 (R Core Team). ChatGPT (OpenAI; https://chatgpt.com) was used to assist with developing and debugging R code and improving the readability and language of the manuscript. The authors reviewed and verified the analyses and retained responsibility for the final manuscript.

## Results

A total of 5 526 599 individuals (2 784 897 [50.4%] women) were followed for 63 million person-years. During follow-up, 1 463 064 (26.5%) received at least one mental, behavioural, or neurodevelopmental disorder diagnosis, including 837 323 (30.1%) women and 625 741 (22.8%) men. SUD was diagnosed in 119 024 (4.3%) men and 58 859 (2.1%) women. Numbers of individuals and mortality rates according to diagnosis, age groups, gender, psychiatric care setting, SUD status, and model covariates are presented in Supplementary Tables S1–S5.

Depressive disorders, anxiety or fear-related disorders, and disorders specifically associated with stress were the three most common diagnostic groups overall (Table 1) and were commonly recorded outside specialist psychiatric care (Supplementary Table S1). Among individuals aged 5–64 years with a mental disorder, 861 842 (75.3%) had at least one of these three disorders. Of these, 527 905 had neither a diagnosed SUD nor specialist psychiatric care during follow-up, accounting for 46.1% of all individuals with a mental disorder in this age group. Among individuals aged 65–95 years, dementia and depressive disorders were the most common diagnostic groups (Supplementary Table S2).

**Table 1.**
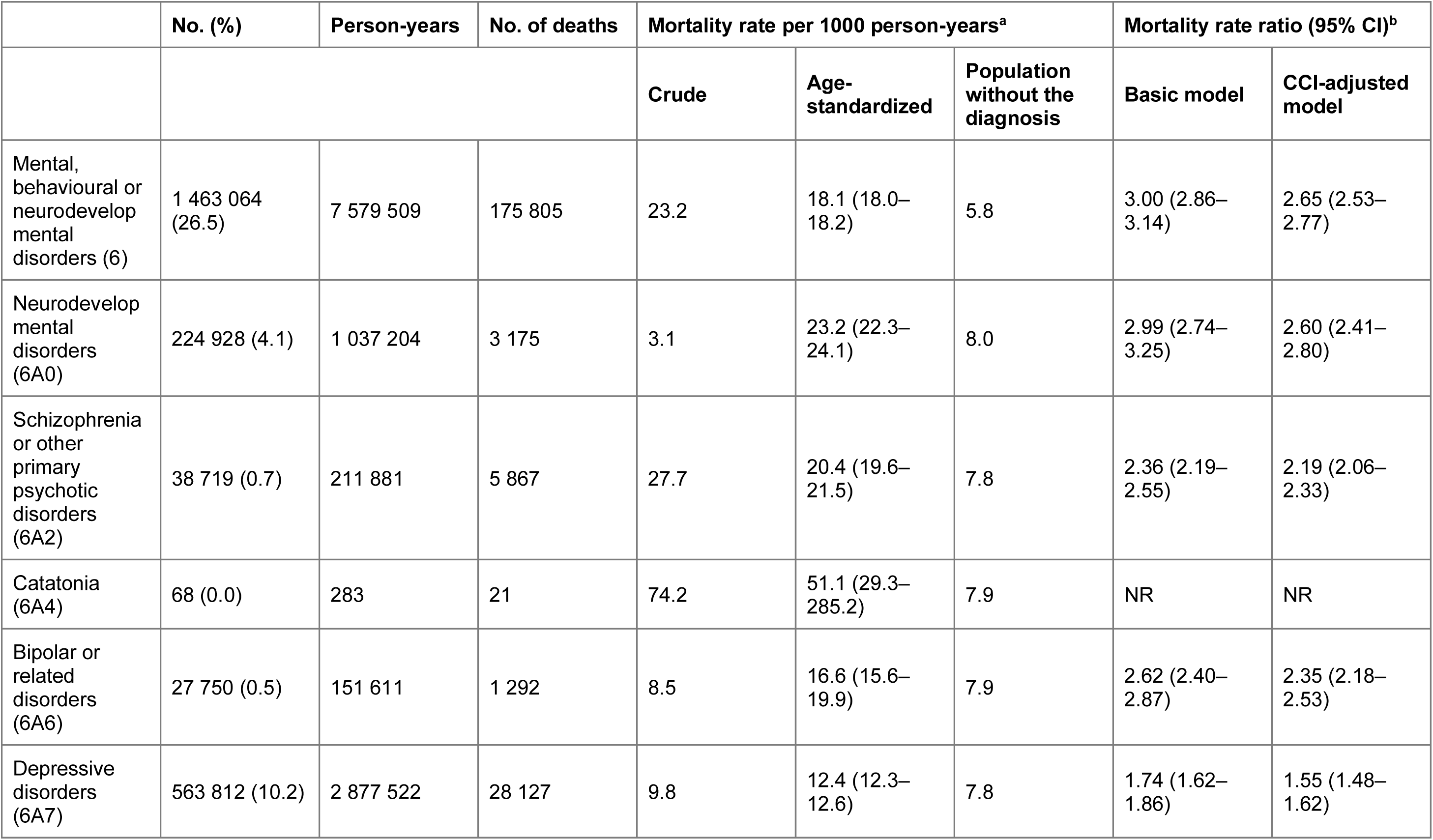
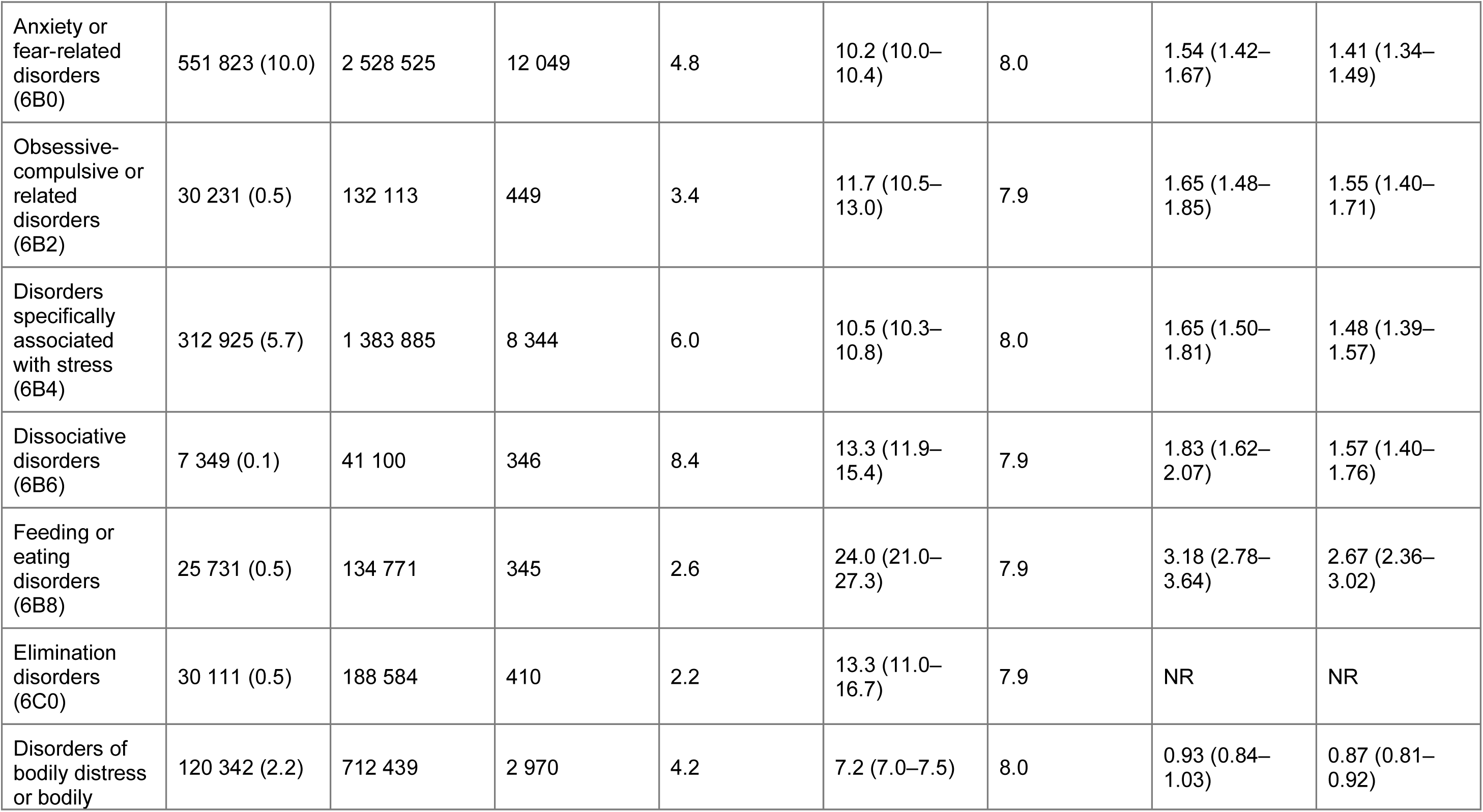
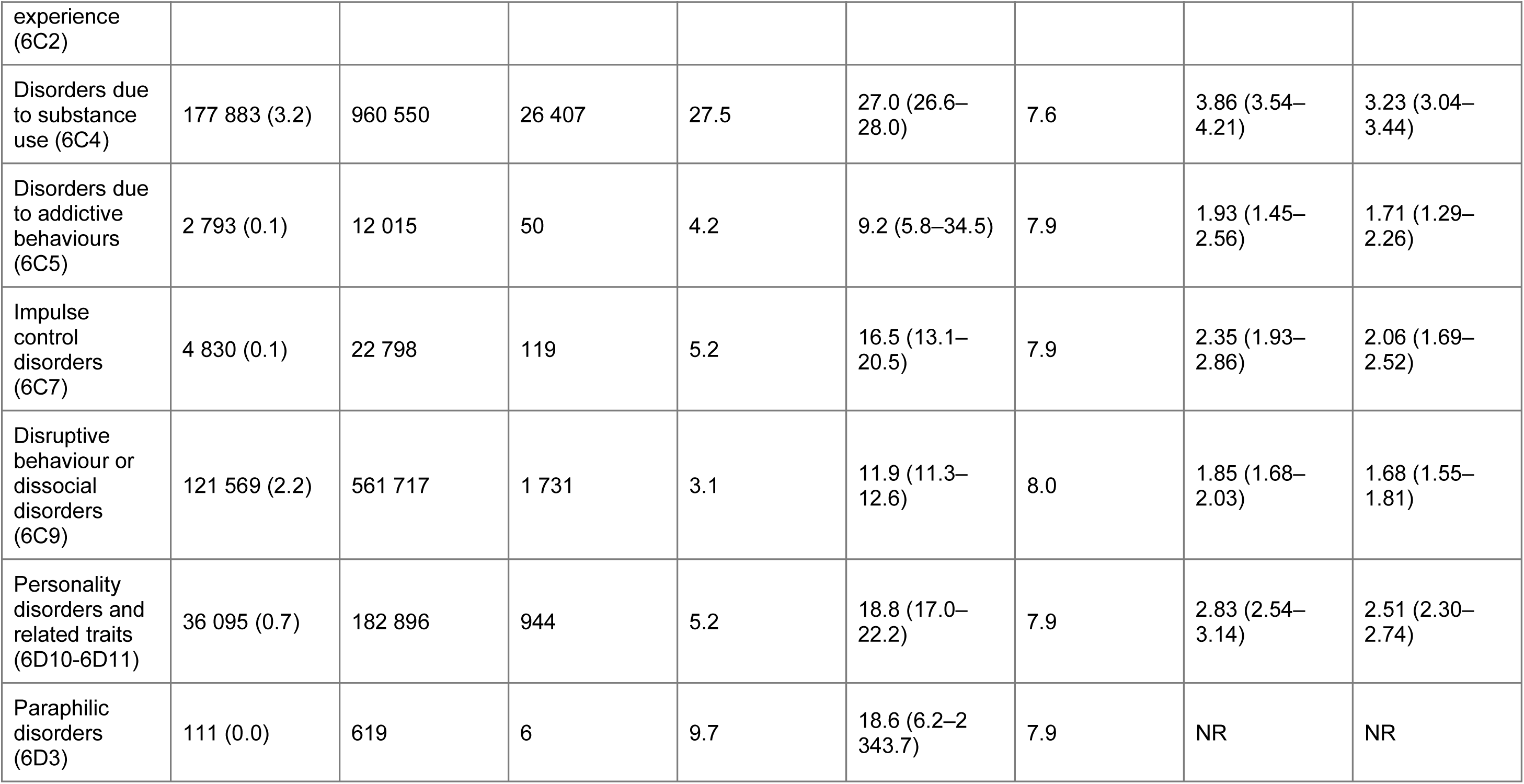
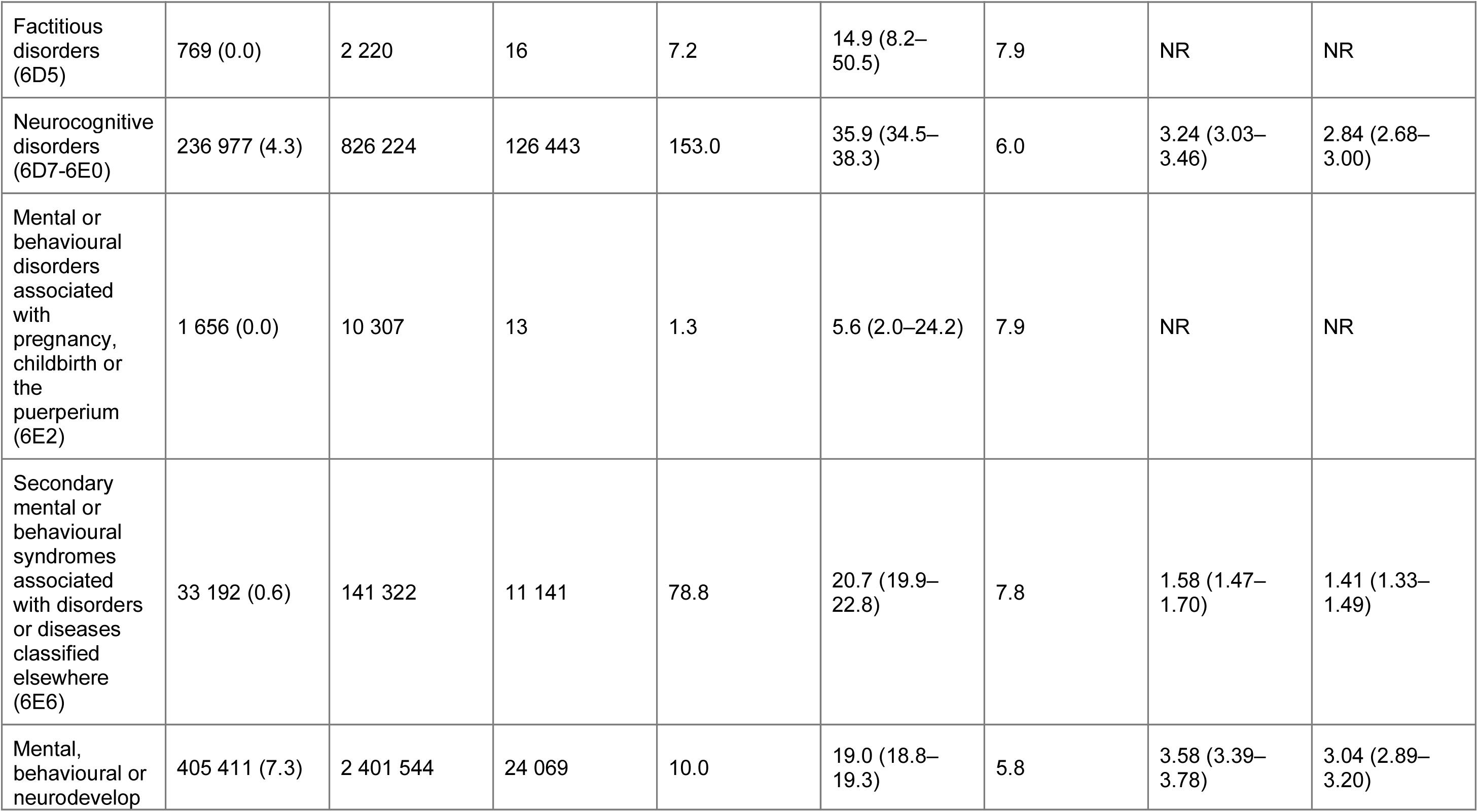
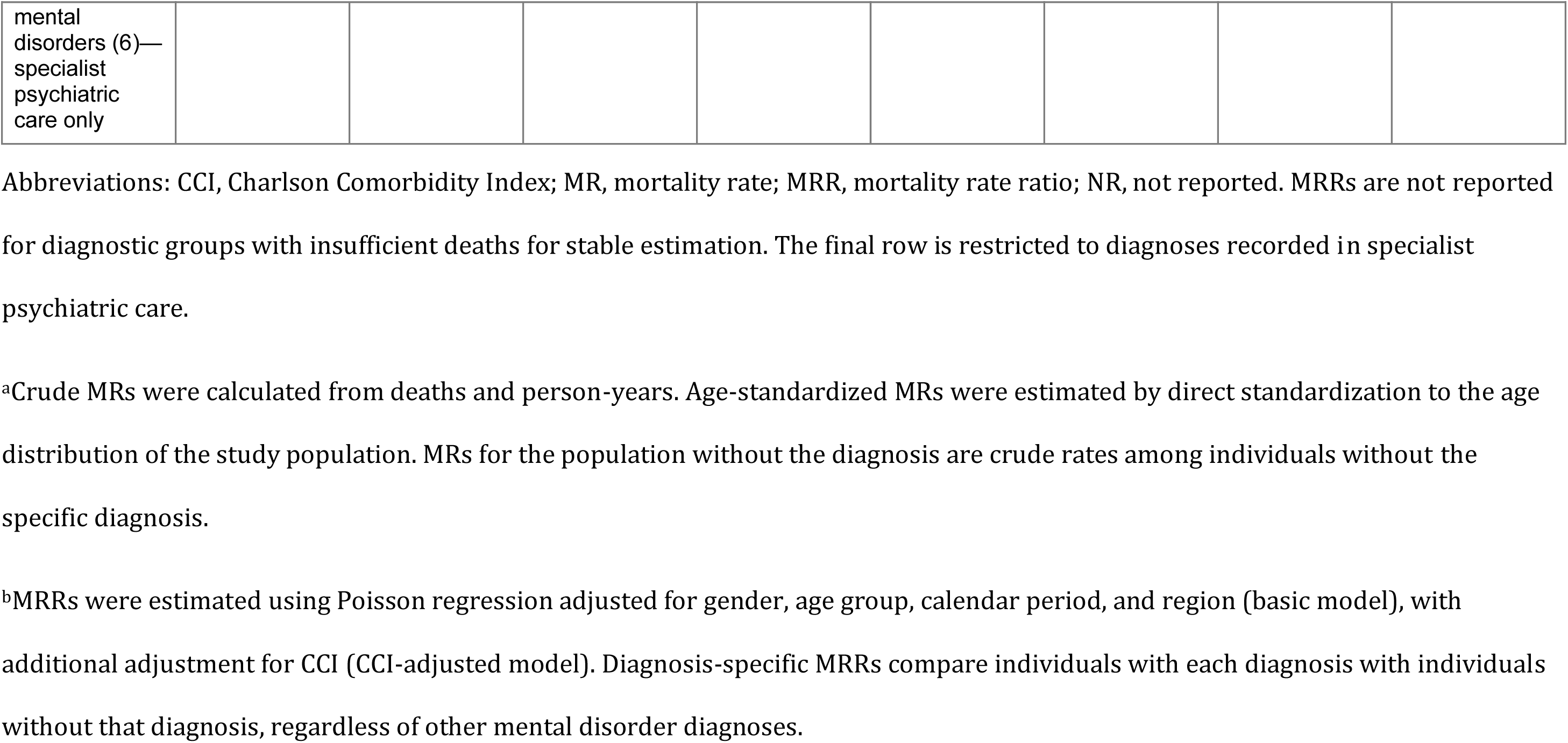
Mortality rates and mortality rate ratios associated with mental, behavioural, and neurodevelopmental disorders.

|  | **No. (%)** | **Person-years** | **No. of deaths** | **Mortality rate per 1000 person-years<sup>a</sup>** |  |  | **Mortality rate ratio (95% CI)<sup>b</sup>** |  |
|---|---|---|---|---|---|---|---|---|
|  |  |  |  | **Crude** | **Age-standardized** | **Population without the diagnosis** | **Basic model** | **CCI-adjusted model** |
| Mental, behavioural or neurodevelopmental disorders (6) | 1 463 064 (26.5) | 7 579 509 | 175 805 | 23.2 | 18.1 (18.0–18.2) | 5.8 | 3.00 (2.86–3.14) | 2.65 (2.53–2.77) |
| Neurodevelopmental disorders (6A0) | 224 928 (4.1) | 1 037 204 | 3 175 | 3.1 | 23.2 (22.3–24.1) | 8.0 | 2.99 (2.74–3.25) | 2.60 (2.41–2.80) |
| Schizophrenia or other primary psychotic disorders (6A2) | 38 719 (0.7) | 211 881 | 5 867 | 27.7 | 20.4 (19.6–21.5) | 7.8 | 2.36 (2.19–2.55) | 2.19 (2.06–2.33) |
| Catatonia (6A4) | 68 (0.0) | 283 | 21 | 74.2 | 51.1 (29.3–285.2) | 7.9 | NR | NR |
| Bipolar or related disorders (6A6) | 27 750 (0.5) | 151 611 | 1 292 | 8.5 | 16.6 (15.6–19.9) | 7.9 | 2.62 (2.40–2.87) | 2.35 (2.18–2.53) |
| Depressive disorders (6A7) | 563 812 (10.2) | 2 877 522 | 28 127 | 9.8 | 12.4 (12.3–12.6) | 7.8 | 1.74 (1.62–1.86) | 1.55 (1.48–1.62) |
| Anxiety or fear-related disorders (6B0) | 551 823 (10.0) | 2 528 525 | 12 049 | 4.8 | 10.2 (10.0–10.4) | 8.0 | 1.54 (1.42–1.67) | 1.41 (1.34–1.49) |
| Obsessive-compulsive or related disorders (6B2) | 30 231 (0.5) | 132 113 | 449 | 3.4 | 11.7 (10.5–13.0) | 7.9 | 1.65 (1.48–1.85) | 1.55 (1.40–1.71) |
| Disorders specifically associated with stress (6B4) | 312 925 (5.7) | 1 383 885 | 8 344 | 6.0 | 10.5 (10.3–10.8) | 8.0 | 1.65 (1.50–1.81) | 1.48 (1.39–1.57) |
| Dissociative disorders (6B6) | 7 349 (0.1) | 41 100 | 346 | 8.4 | 13.3 (11.9–15.4) | 7.9 | 1.83 (1.62–2.07) | 1.57 (1.40–1.76) |
| Feeding or eating disorders (6B8) | 25 731 (0.5) | 134 771 | 345 | 2.6 | 24.0 (21.0–27.3) | 7.9 | 3.18 (2.78–3.64) | 2.67 (2.36–3.02) |
| Elimination disorders (6C0) | 30 111 (0.5) | 188 584 | 410 | 2.2 | 13.3 (11.0–16.7) | 7.9 | NR | NR |
| Disorders of bodily distress or bodily experience (6C2) | 120 342 (2.2) | 712 439 | 2 970 | 4.2 | 7.2 (7.0–7.5) | 8.0 | 0.93 (0.84–1.03) | 0.87 (0.81–0.92) |
| Disorders due to substance use (6C4) | 177 883 (3.2) | 960 550 | 26 407 | 27.5 | 27.0 (26.6–28.0) | 7.6 | 3.86 (3.54–4.21) | 3.23 (3.04–3.44) |
| Disorders due to addictive behaviours (6C5) | 2 793 (0.1) | 12 015 | 50 | 4.2 | 9.2 (5.8–34.5) | 7.9 | 1.93 (1.45–2.56) | 1.71 (1.29–2.26) |
| Impulse control disorders (6C7) | 4 830 (0.1) | 22 798 | 119 | 5.2 | 16.5 (13.1–20.5) | 7.9 | 2.35 (1.93–2.86) | 2.06 (1.69–2.52) |
| Disruptive behaviour or dissocial disorders (6C9) | 121 569 (2.2) | 561 717 | 1 731 | 3.1 | 11.9 (11.3–12.6) | 8.0 | 1.85 (1.68–2.03) | 1.68 (1.55–1.81) |
| Personality disorders and related traits (6D10-6D11) | 36 095 (0.7) | 182 896 | 944 | 5.2 | 18.8 (17.0–22.2) | 7.9 | 2.83 (2.54–3.14) | 2.51 (2.30–2.74) |
| Paraphilic disorders (6D3) | 111 (0.0) | 619 | 6 | 9.7 | 18.6 (6.2–2343.7) | 7.9 | NR | NR |
| Factitious disorders (6D5) | 769 (0.0) | 2 220 | 16 | 7.2 | 14.9 (8.2–50.5) | 7.9 | NR | NR |
| Neurocognitive disorders (6D7-6E0) | 236 977 (4.3) | 826 224 | 126 443 | 153.0 | 35.9 (34.5–38.3) | 6.0 | 3.24 (3.03–3.46) | 2.84 (2.68–3.00) |
| Mental or behavioural disorders associated with pregnancy, childbirth or the puerperium (6E2) | 1 656 (0.0) | 10 307 | 13 | 1.3 | 5.6 (2.0–24.2) | 7.9 | NR | NR |
| Secondary mental or behavioural syndromes associated with disorders or diseases classified elsewhere (6E6) | 33 192 (0.6) | 141 322 | 11 141 | 78.8 | 20.7 (19.9–22.8) | 7.8 | 1.58 (1.47–1.70) | 1.41 (1.33–1.49) |
| Mental, behavioural or neurodevelopmental disorders (6)—specialist psychiatric care only | 405 411 (7.3) | 2 401 544 | 24 069 | 10.0 | 19.0 (18.8–19.3) | 5.8 | 3.58 (3.39–3.78) | 3.04 (2.89–3.20) |

Abbreviations: CCI, Charlson Comorbidity Index; MR, mortality rate; MRR, mortality rate ratio; NR, not reported. MRRs are not reported for diagnostic groups with insufficient deaths for stable estimation. The final row is restricted to diagnoses recorded in specialist psychiatric care.
<sup>a</sup>Crude MRs were calculated from deaths and person-years. Age-standardized MRs were estimated by direct standardization to the age distribution of the study population. MRs for the population without the diagnosis are crude rates among individuals without the specific diagnosis.
<sup>b</sup>MRRs were estimated using Poisson regression adjusted for gender, age group, calendar period, and region (basic model), with additional adjustment for CCI (CCI-adjusted model). Diagnosis-specific MRRs compare individuals with each diagnosis with individuals without that diagnosis, regardless of other mental disorder diagnoses.

Excess mortality varied across ICD-11 diagnostic groups (Table 1). Adjustment for baseline physical comorbidity reduced MRRs, but substantial variation across diagnostic groups remained. Among diagnostic groups with stable estimates, SUD had the highest adjusted MRR (3.23, 95% CI, 3.04–3.44), whereas MRRs were lower for depressive disorders (1.55, 1.48–1.62), anxiety or fear-related disorders (1.41, 1.34–1.49), and disorders specifically associated with stress (1.48, 1.39–1.57).

MRRs also varied substantially with age and case ascertainment (Figure 1). Restricting diagnoses to specialist psychiatric care produced higher MRRs at younger ages but lower MRRs at older ages than ascertainment from both primary and specialist care.

**Figure 1.**
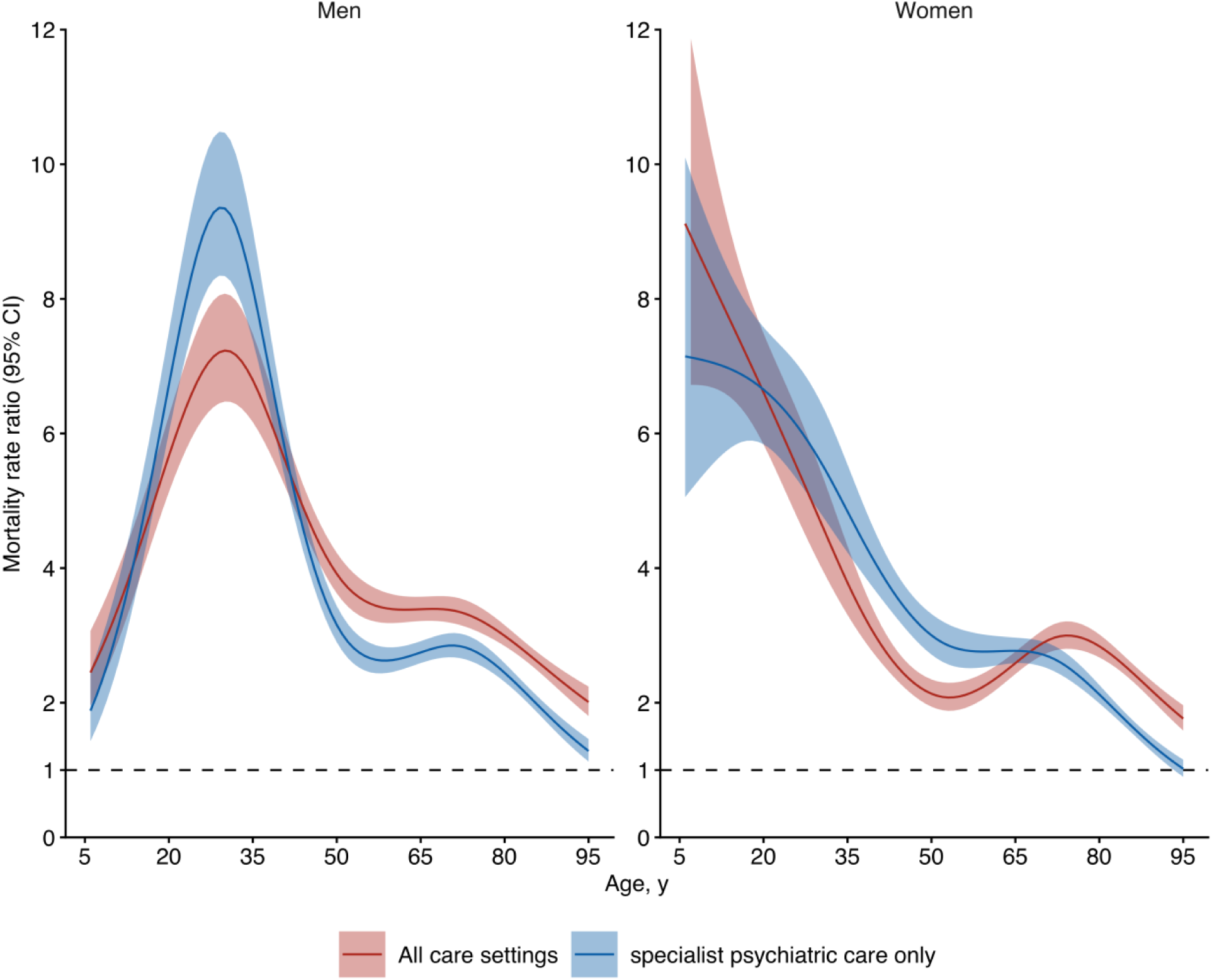
Age-specific mortality rate ratios for mental, behavioural, and neurodevelopmental disorders by case ascertainment. MRRs are relative to individuals without a diagnosed mental, behavioural, or neurodevelopmental disorder. The all care settings analysis included diagnoses recorded both within and outside specialist psychiatric care. In the specialist psychiatric care only analysis, individuals diagnosed exclusively outside specialist psychiatric care were classified as having no mental disorder diagnosis. MRRs were estimated separately for women and men using Poisson regression adjusted for calendar period, region, and Charlson Comorbidity Index. Shaded areas indicate 95% CIs; the dashed horizontal line indicates an MRR of 1.

Across all mental, behavioural, and neurodevelopmental disorders combined, MRRs were highest immediately after diagnosis and declined during the first two years across all clinical subgroups (Figure 2). Excess mortality nevertheless remained elevated overall throughout follow-up, with the highest MRRs among individuals with SUD receiving specialist psychiatric care and the lowest among those without SUD treated outside specialist psychiatric care. Temporal patterns were similar for natural and external causes, although relative mortality was higher for external causes, particularly among individuals with SUD receiving specialist psychiatric care. Follow-up-averaged MRRs according to diagnostic group, SUD status, and psychiatric care setting are presented separately for individuals aged 5–64 and 65–95 years in Supplementary Tables S6 and S7.

**Figure 2.**
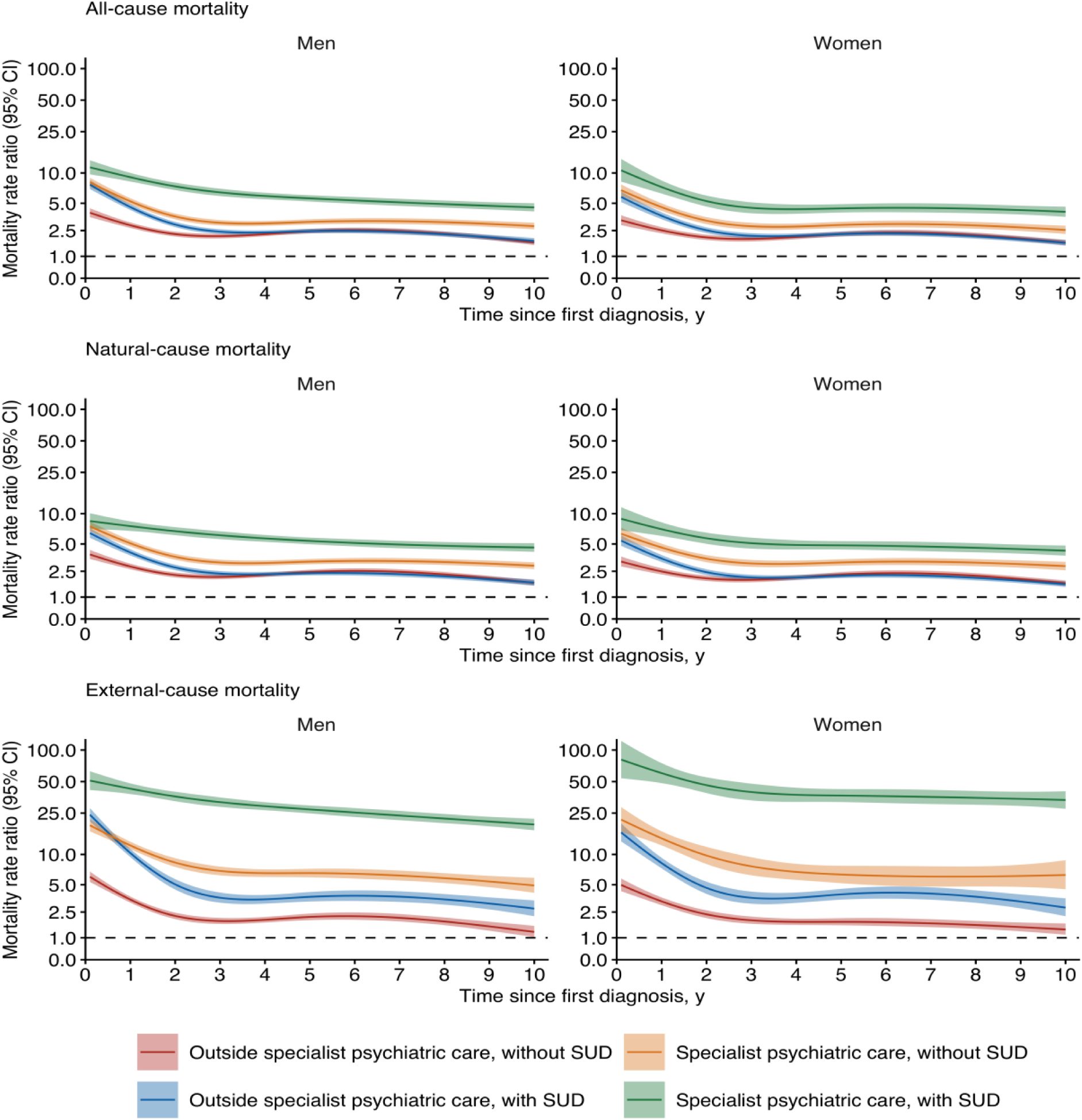
Mortality rate ratios over time since first diagnosis of mental disorders by psychiatric care setting, substance use disorder status, and cause of death. Abbreviations: MRR, mortality rate ratio; SUD, substance use disorder. MRRs are relative to individuals without a diagnosed mental, behavioural, or neurodevelopmental disorder. Person-time was dynamically allocated according to time-varying psychiatric care setting and SUD status. MRRs were estimated separately for women and men using Poisson regression adjusted for age group, calendar period, region, and Charlson Comorbidity Index. Estimates are shown for the first 10 years after diagnosis. Shaded areas indicate 95% CIs; the dashed horizontal line indicates an MRR of 1. The y-axis is logarithmic.

Mortality trajectories varied substantially not only across diagnostic groups but also within diagnostic groups according to SUD status, psychiatric care setting, and age (Figures 3 and 4). Among individuals aged 5–64 years with depressive, anxiety or fear-related, or stress-related disorders who had no diagnosed SUD and were treated outside specialist psychiatric care—46.1% of all individuals with a mental disorder in this age group—MRRs declined rapidly after diagnosis. At two years, MRRs ranged from 0.88 (95% CI 0.73–1.06) to 1.04 (0.88–1.22) in men and from 0.92 (0.79–1.07) to 1.12 (0.96–1.31) in women across these three diagnostic groups, with little evidence of excess mortality thereafter. For these disorders, excess mortality was more persistent among individuals receiving specialist psychiatric care, whereas among those with SUD treated outside specialist psychiatric care it declined substantially over time. Similar temporal patterns were observed among individuals aged 65–95 years, although for depressive disorders excess mortality persisted across all clinical subgroups. Schizophrenia and other primary psychotic disorders showed a different pattern, with excess mortality persisting even among individuals without SUD treated outside specialist psychiatric care. Time-varying MRRs for the remaining diagnostic groups, including cause-specific estimates, are presented in Supplementary Figures S1–S14.

**Figure 3.**
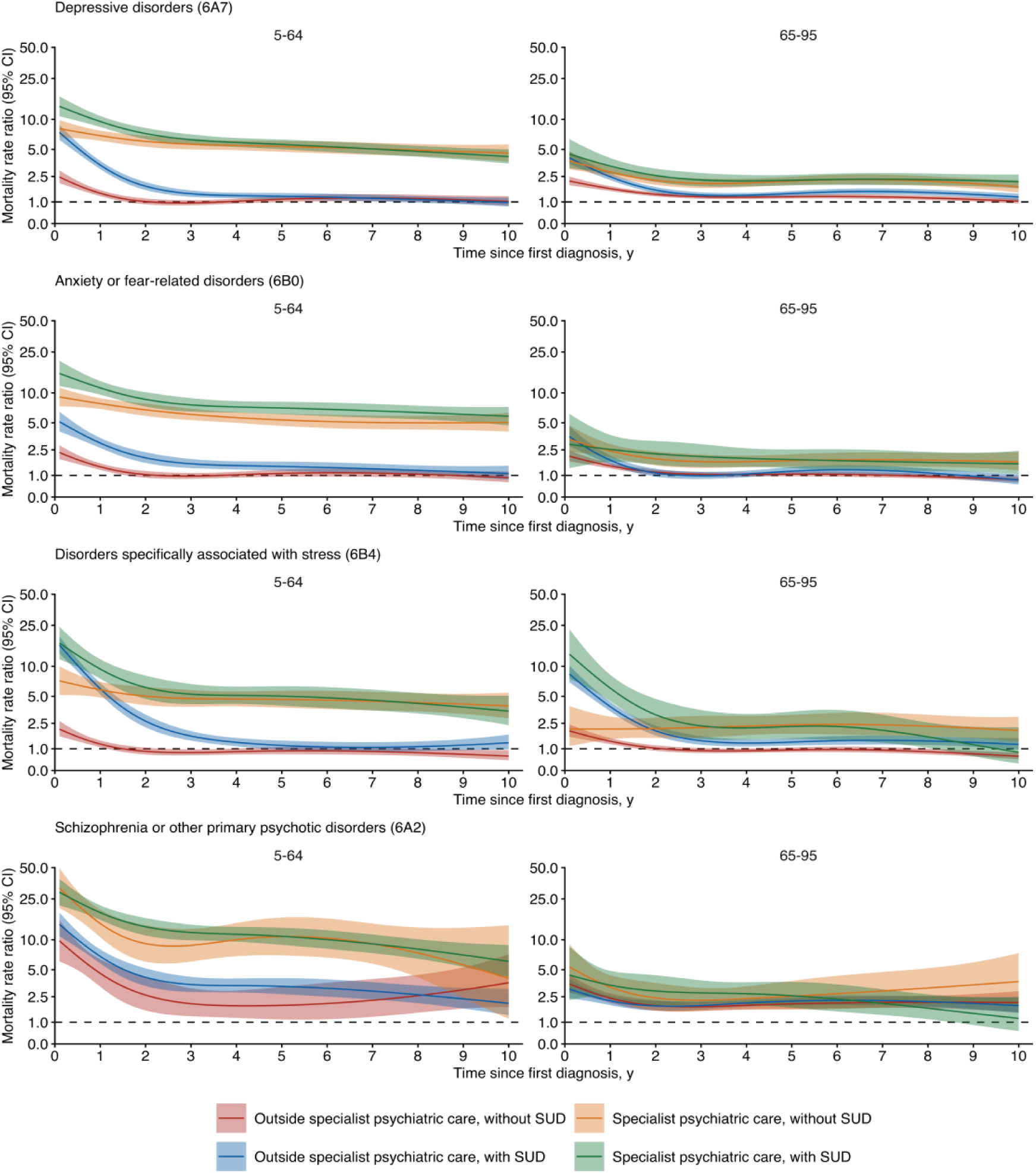
Mortality rate ratios over time since first diagnosis for selected mental disorders by psychiatric care setting, substance use disorder status, and age group in men. Abbreviations: MRR, mortality rate ratio; SUD, substance use disorder. MRRs are relative to individuals without the specific diagnostic group. Person-time was dynamically allocated according to time-varying psychiatric care setting and SUD status. MRRs were estimated using Poisson regression adjusted for age group, calendar period, region, and Charlson Comorbidity Index. Estimates are shown for the first 10 years after diagnosis. Shaded areas indicate 95% CIs; the dashed horizontal line indicates an MRR of 1. The y-axis is logarithmic.

**Figure 4.**
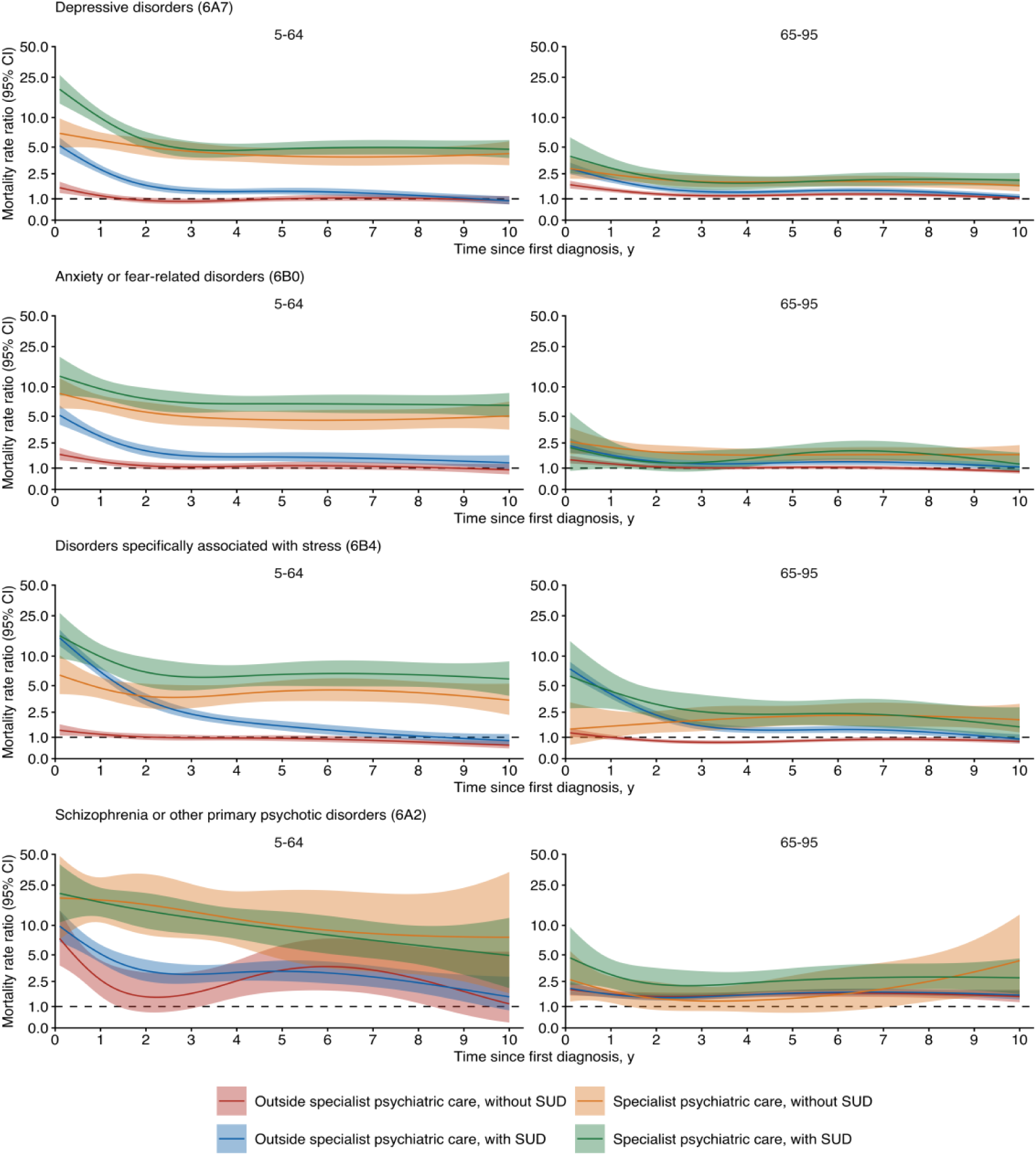
Mortality rate ratios over time since first diagnosis for selected mental disorders by psychiatric care setting, substance use disorder status, and age group in women. Abbreviations: MRR, mortality rate ratio; SUD, substance use disorder. MRRs are relative to individuals without the specific diagnostic group. Person-time was dynamically allocated according to time-varying psychiatric care setting and SUD status. MRRs were estimated separately using Poisson regression adjusted for age group, calendar period, region, and Charlson Comorbidity Index. Estimates are shown for the first 10 years after diagnosis. Shaded areas indicate 95% CIs; the dashed horizontal line indicates an MRR of 1. The y-axis is logarithmic.

Differences in relative mortality were also reflected in absolute survival (Supplementary Tables S8 and S9). Among individuals without SUD treated outside specialist psychiatric care, ten-year survival loss was small for depressive disorders (0.09 years in men and 0.05 in women), anxiety or fear-related disorders (0.03 and 0.01 years, respectively), and stress-related disorders (0.01 and −0.01 years). Thus, for these common disorders, estimated survival loss in this clinical group was generally measured in days rather than months. Among individuals with SUD receiving specialist psychiatric care, the corresponding differences were substantially larger: 0.67 years in men and 0.43 years in women for depressive disorders, 0.45 and 0.27 years, respectively, for anxiety or fear-related disorders, and 0.59 and 0.39 for stress-related disorders. Across all mental, behavioural, and neurodevelopmental disorders, ten-year survival loss was 0.53 years in men and 0.34 years in women, with survival loss associated with natural causes substantially greater than that associated with external causes of death. The corresponding RMST estimates for individuals with and without each diagnostic group are presented in Supplementary Tables S10 and S11.

Results were similar when analyses were restricted to disorders first diagnosed from 2016 onward (Figure S15). Using ICD-10 main diagnostic groups resulted in more individuals being classified as having a mental disorder than using the ICD-11 grouping (32.0% vs 26.5%) and yielded lower MRRs for mental disorders combined, particularly when diagnoses outside specialist psychiatric care were included; estimates based solely on specialist psychiatric care were similar (Supplementary Tables S12 and S13).

## Discussion

In this nationwide study, excess mortality associated with mental disorders differed markedly according to clinical context and time since diagnosis. Among individuals aged 5–64 years, those with depressive, anxiety or fear-related, or stress-related disorders who had no diagnosed SUD and did not receive specialist psychiatric care during follow-up accounted for over 46% of all individuals with a mental disorder. In this large group, mortality was elevated immediately after diagnosis but declined rapidly during the first two years, with little evidence of excess mortality thereafter. In contrast, excess mortality was more persistent among individuals receiving specialist psychiatric care and particularly high after diagnosis among those with SUD. Schizophrenia and other primary psychotic disorders showed a different pattern, with excess mortality persisting even among individuals without SUD treated outside specialist psychiatric care. A single follow-up-averaged mortality estimate for a diagnostic group therefore obscures a central feature of the mortality gap: excess mortality may be transient in a large part of the diagnosed population while remaining substantial or persistent in others with the same diagnosis. Distinguishing when and in whom excess mortality occurs is important for better targeting preventive efforts and for giving patients and clinicians a more informative account of how mortality risk changes after diagnosis.

Previous studies have shown that excess mortality following depression, anxiety, and stress-related disorders is highest shortly after diagnosis and declines with time, although studies based on secondary psychiatric care have found excess mortality persisting during longer follow-up [16–18]. Recent meta-analyses nevertheless show substantial variation according to the populations studied [8,9]. For depression, mortality remains elevated in community samples, whereas an earlier meta-analysis found no excess mortality associated with anxiety in community samples [8,19]. Our findings bring these dimensions together by showing that the temporal pattern itself differs according to clinical context. The persistent excess mortality observed in specialist psychiatric populations should therefore not be assumed to characterize the broader—and substantially larger— population with common mental disorders diagnosed outside specialist psychiatric care. This difference also has implications for case ascertainment in mortality studies. Previous studies have shown that mortality estimates based on specialist psychiatric populations may differ substantially from those in the broader population [10,20,21]. Our findings further show that the implications of restricting ascertainment to specialist psychiatric care vary across the life course. Such restriction produced particularly high relative mortality estimates in young and middle-aged adults, but lower estimates at older ages than ascertainment from both primary and specialist care. Thus, specialist psychiatric populations do not represent a uniformly higher-mortality subset of people with mental disorders, and their relationship to the broader diagnosed population changes with age.

Age also modified the temporal patterns within diagnostic groups. Among individuals aged 65–95 years, depression was an important exception to the attenuation observed for common mental disorders at younger ages: excess mortality persisted across clinical subgroups, including among those without SUD treated outside specialist psychiatric care. A similar persistent pattern was not observed for anxiety or fear-related disorders. Previous studies have consistently associated late-life depression with increased mortality in community populations [22]. Notably, in our study excess mortality persisted after adjustment for baseline physical comorbidity. This distinct mortality trajectory is consistent with broader evidence that depression in later life differs from depression in younger adults in its clinical characteristics, prognosis, and relationships with physical and neurocognitive disorders [23].

SUD was an important marker of mortality within diagnostic groups, consistent with the substantial excess mortality associated with SUD and co-occurring mental disorders reported previously [11,24–27]. However, SUD status alone did not distinguish transient from persistent excess mortality. Among individuals with common mental disorders, excess mortality declined substantially among those with SUD treated outside specialist psychiatric care, whereas it remained elevated among those receiving specialist psychiatric care even without SUD. Treating SUD solely as a covariate may therefore obscure clinically important heterogeneity, as mortality trajectories differed substantially according to SUD status, clinical setting, and time since diagnosis even within the same diagnostic group. These differences were also apparent on an absolute scale, with substantially greater ten-year survival loss among individuals with SUD and those receiving specialist psychiatric care than among those without SUD treated outside specialist psychiatric care.

Diagnostic classification provided another source of variation in mortality estimates. Applying the ICD-11 framework classified fewer individuals as having a mental, behavioural, or neurodevelopmental disorder than ICD-10 main diagnostic groups and yielded higher overall MRRs, particularly when diagnoses outside specialist psychiatric care were included. Thus, diagnostic classification can change both the population identified as having a mental disorder and the resulting estimate of the mortality gap.

This study has several limitations. First, primary care data were available only from 2011, and some prevalent disorders may therefore have been misclassified as incident, although sensitivity analyses restricted to more recent diagnoses yielded similar results. Second, ICD-11 diagnostic groups were derived from ICD-10 codes using WHO mapping tables rather than diagnoses recorded using ICD-11. Third, SUDs are likely to be underascertained in health care registers, which may have reduced observed differences according to SUD status. SUD status and psychiatric care setting should be interpreted as markers of clinical context rather than causal determinants of mortality; both may reflect disorder severity, other patient characteristics, and patterns of health care contact. Fourth, time since diagnosis refers to time since the first recorded diagnosis. The analyses could not establish when a disorder began or distinguish active disorder, remission, and recurrence, and the observed temporal patterns should therefore not be interpreted as effects of diagnosis itself. Fifth, physical comorbidity was assessed at cohort entry and not updated during follow-up, so subsequent changes in physical health were not captured by CCI adjustment. Finally, the ten-year RMST differences quantify survival lost during the first ten years after diagnosis and should not be interpreted as reductions in life expectancy or extrapolated beyond the observed follow-up.

In conclusion, the mortality gap associated with mental disorders cannot be adequately characterized by diagnostic group or a single summary estimate. Excess mortality may be transient in large parts of the diagnosed population while remaining substantial or persistent in others with the same diagnosis, depending on time since diagnosis and clinical context. Recognizing this heterogeneity may provide a stronger basis for targeting prevention and for giving patients and clinicians a more informative account of mortality after diagnosis.

## Data Availability

The data used in this study are maintained by Statistics Finland and the Finnish Institute for Health and Welfare (THL), which act as data controllers for the respective datasets. Access to the data is subject to restrictions and requires authorization from the relevant authorities. For more information, see www.findata.fi and www.stat.fi.

## Supplementary material

For supplementary material accompanying this paper, visit cambridge.org/EPA.

