## Supplementary material for "Excess mortality after mental disorder diagnosis within and outside specialist psychiatric care: nationwide cohort study"

### Table of contents

Supplementary methods ... 3

Table S1. Diagnosis-specific mortality rates by psychiatric care setting ... 4

Table S2. Diagnosis-specific mortality rates by age group ... 7

Table S3. Diagnosis-specific mortality rates by gender ... 10

Table S4. Diagnosis-specific mortality rates by substance use disorder status ... 13

Table S5. Mortality rates by regression model covariates ... 16

Table S6. Diagnosis-specific mortality rate ratios by psychiatric care setting and substance use disorder status among individuals aged 5 to 64 years ... 18

Table S7. Diagnosis-specific mortality rate ratios by psychiatric care setting and substance use disorder status among individuals aged 65 to 95 years ... 21

Figure S1. Time-varying mortality rate ratios for neurodevelopmental disorders (6A0) by psychiatric care setting, substance use disorder status, and cause of death ... 24

Figure S2. Time-varying mortality rate ratios for schizophrenia or other primary psychotic disorders (6A2) by psychiatric care setting, substance use disorder status, and cause of death ... 25

Figure S3. Time-varying mortality rate ratios for bipolar or related disorders (6A6) by psychiatric care setting, substance use disorder status, and cause of death ... 26

Figure S4. Time-varying mortality rate ratios for depressive disorders (6A7) by psychiatric care setting, substance use disorder status, and cause of death ... 27

Figure S5. Time-varying mortality rate ratios for anxiety or fear-related disorders (6B0) by psychiatric care setting, substance use disorder status, and cause of death ... 28

Figure S6. Time-varying mortality rate ratios for obsessive-compulsive or related disorders (6B2) by psychiatric care setting, substance use disorder status, and cause of death ... 29

Figure S7. Time-varying mortality rate ratios for disorders specifically associated with stress (6B4) by psychiatric care setting, substance use disorder status, and cause of death ... 30

Figure S8. Time-varying mortality rate ratios for dissociative disorders (6B6) by psychiatric care setting, substance use disorder status, and cause of death ... 31

Figure S9. Time-varying mortality rate ratios for feeding or eating disorders (6B8) by psychiatric care setting, substance use disorder status, and cause of death ... 32

Figure S10. Time-varying mortality rate ratios for disorders of bodily distress or bodily experience (6C2) by psychiatric care setting, substance use disorder status, and cause of death ... 33

Figure S11. Time-varying mortality rate ratios for personality disorders and related traits (6D10-6D11) by psychiatric care setting, substance use disorder status, and cause of death ... 34

Figure S12. Time-varying mortality rate ratios for neurocognitive disorders (6D7-6E0) by psychiatric care setting, substance use disorder status, and cause of death ... 35

Figure S13. Time-varying mortality rate ratios for dementia (6D8) by psychiatric care setting, substance use disorder status, and cause of death ... 36

Figure S14. Time-varying mortality rate ratios for secondary mental or behavioural syndromes associated with disorders or diseases classified elsewhere (6E6) by psychiatric care setting, substance use disorder status, and cause of death ... 37

**Supplementary methods**

ICD-10 has been used in Finland since 1996; Finnish versions of ICD-9 (1987–1995) and ICD-8 (1969–1986) were used previously. Some primary care facilities record diagnoses using the International Classification of Primary Care, Second Edition (ICPC-2). ICD-8, ICD-9, and ICPC-2 diagnoses were converted to corresponding ICD-10 subchapter categories. Register data were preprocessed to maximize diagnostic consistency across data sources and classification systems. Preprocessing included harmonization of overlapping and transfer-related register entries, timing of hospital episodes, and exclusion of preliminary diagnoses.

ICD-10-to-ICD-11 mappings were based on the WHO mapping tables downloaded from cdcdn.who.int/static/releasefiles/2025-01/mapping.zip on May 22, 2025. To ensure mutually exclusive ICD-11 diagnostic groupings, we used the 10to11MapToOneCategory mappings rather than multiple-category mappings.

Three manual modifications were made to improve clinical specificity.

1. Manic episode diagnoses (F30.0–F30.9) were assigned to bipolar or related disorders (6A6Z), consistent with ICD-11 diagnostic requirements whereby a history of a single manic episode is sufficient for bipolar type I disorder.

2. Puerperal mental and behavioural disorders (F53.0–F53.9) were assigned to mental or behavioural disorders associated with pregnancy, childbirth or the puerperium (6E2Z) rather than to the unspecified mental, behavioural or neurodevelopmental disorder category (6E8Z).

3. F84.8 was assigned to autism spectrum disorder (6A02.Z), reflecting Finnish clinical coding practice whereby Asperger's syndrome has commonly been recorded using F84.8 pending ICD-11 implementation.

**Table S1. Diagnosis-specific mortality rates by psychiatric care setting**

| | Outside specialist psychiatric care | | | | | Specialist psychiatric care | | | | |
|---|---|---|---|---|---|---|---|---|---|---|
| | No. (%) | Person-years | No. of deaths | Mortality rate per 1000 person-years | | No. (%) | Person-years | No. of deaths | Mortality rate per 1000 person-years | |
| | | | | Crude | Age-standardized | | | | Crude | Age-standardized |
| Mental, behavioural or neurodevelopmental disorders (6) | 1 265 893 (22.9) | 5 177 965 | 151 736 | 29.3 | 17.7 (17.57–17.77) | 405 411 (7.3) | 2 401 544 | 24 069 | 10.0 | 19.0 (18.78–19.28) |
| Neurodevelopmental disorders (6A0) | 171 296 (3.1) | 677 717 | 2 779 | 4.1 | 24.2 (23.30–25.23) | 83 982 (1.5) | 359 487 | 396 | 1.1 | 22.6 (12.78–40.83) |
| Schizophrenia or other primary psychotic disorders (6A2) | 14 167 (0.3) | 40 740 | 2 309 | 56.7 | 22.9 (21.41–25.11) | 30 065 (0.5) | 171 141 | 3 558 | 20.8 | 19.2 (18.54–21.00) |
| Catatonia (6A4) | 46 (0.0) | 137 | 16 | 116.7 | 75.1 (34.98–313.00) | 32 (0.0) | NR | NR | NR | NR |
| Bipolar or related disorders (6A6) | 12 338 (0.2) | 43 225 | 576 | 13.3 | 17.1 (15.64–26.91) | 19 451 (0.4) | 108 387 | 716 | 6.6 | 15.2 (13.60–20.16) |
| Depressive disorders (6A7) | 435 691 (7.9) | 1 566 478 | 18 572 | 11.9 | 11.4 (11.24–11.86) | 221 813 (4.0) | 1 311 044 | 9 555 | 7.3 | 14.5 (14.17–14.87) |
| Anxiety or fear-related disorders (6B0) | 437 005 (7.9) | 1 559 693 | 8 369 | 5.4 | 9.5 (9.31–9.73) | 180 173 (3.3) | 968 832 | 3 680 | 3.8 | 12.3 (11.83–12.78) |
| Obsessive-compulsive or related disorders (6B2) | 17 481 (0.3) | 50 632 | 282 | 5.6 | 11.7 (10.33–13.31) | 16 339 (0.3) | 81 481 | 167 | 2.0 | 12.4 (9.69–15.79) |
| Disorders specifically associated with stress (6B4) | 260 537 (4.7) | 993 592 | 4 339 | 4.4 | 7.6 (7.38–7.96) | 64 135 (1.2) | 390 293 | 4 005 | 10.3 | 20.3 (19.56–20.98) |

| | Outside specialist psychiatric care | | | | | Specialist psychiatric care | | | | |
|---|---|---|---|---|---|---|---|---|---|---|
| | No. (%) | Person-years | No. of deaths | Mortality rate per 1000 person-years | | No. (%) | Person-years | No. of deaths | Mortality rate per 1000 person-years | |
| | | | | Crude | Age-standardized | | | | Crude | Age-standardized |
| Dissociative disorders (6B6) | 3 849 (0.1) | 18 284 | 264 | 14.4 | 12.8 (11.28–17.13) | 4 045 (0.1) | 22 816 | 82 | 3.6 | 15.6 (11.07–22.16) |
| Feeding or eating disorders (6B8) | 15 269 (0.3) | 40 917 | 239 | 5.8 | 26.5 (23.03–30.40) | 16 930 (0.3) | 93 854 | 106 | 1.1 | 13.4 (5.46–30.31) |
| Elimination disorders (6C0) | 29 492 (0.5) | 179 683 | 407 | 2.3 | 13.3 (10.97–16.71) | 1 561 (0.0) | NR | NR | NR | NR |
| Disorders of bodily distress or bodily experience (6C2) | 117 869 (2.1) | 692 315 | 2 796 | 4.0 | 7.0 (6.76–7.36) | 3 443 (0.1) | 20 125 | 174 | 8.6 | 13.8 (11.70–17.28) |
| Disorders due to substance use (6C4) | 157 072 (2.8) | 758 475 | 22 846 | 30.1 | 26.5 (26.10–27.51) | 33 596 (0.6) | 202 075 | 3 561 | 17.6 | 26.7 (25.51–49.28) |
| Disorders due to addictive behaviours (6C5) | 1 894 (0.0) | 6 297 | 25 | 4.0 | 8.2 (4.90–65.52) | 1 108 (0.0) | 5 718 | 25 | 4.4 | 14.0 (3.28–65.55) |
| Impulse control disorders (6C7) | 3 319 (0.1) | 12 442 | 79 | 6.3 | 16.5 (12.76–21.09) | 1 878 (0.0) | 10 356 | 40 | 3.9 | 18.2 (9.04–34.58) |
| Disruptive behaviour or dissocial disorders (6C9) | 66 405 (1.2) | 204 196 | 1 176 | 5.8 | 11.8 (11.12–12.56) | 64 027 (1.2) | 357 521 | 555 | 1.6 | 11.9 (10.51–13.53) |
| Personality disorders and related traits (6D10-6D11) | 14 631 (0.3) | 53 543 | 371 | 6.9 | 20.0 (17.60–26.35) | 24 247 (0.4) | 129 353 | 573 | 4.4 | 17.1 (14.42–22.92) |
| Paraphilic disorders (6D3) | 57 (0.0) | NR | NR | NR | NR | 62 (0.0) | NR | NR | NR | NR |
| Factitious disorders (6D5) | 593 (0.0) | 1 419 | 13 | 9.2 | 22.3 (11.04–60.47) | 199 (0.0) | NR | NR | NR | NR |

| | Outside specialist psychiatric care | | | | | Specialist psychiatric care | | | | |
|---|---|---|---|---|---|---|---|---|---|---|
| | No. (%) | Person-years | No. of deaths | Mortality rate per 1000 person-years | | No. (%) | Person-years | No. of deaths | Mortality rate per 1000 person-years | |
| | | | | Crude | Age-standardized | | | | Crude | Age-standardized |
| Neurocognitive disorders (6D7-6E0) | 232 065 (4.2) | 782 916 | 119 886 | 153.1 | 34.8 (33.33–37.34) | 11 457 (0.2) | 43 308 | 6 557 | 151.4 | 46.3 (43.80–76.40) |
| Mental or behavioural disorders associated with pregnancy, childbirth or the puerperium (6E2) | 1 174 (0.0) | 6 127 | 9 | 1.5 | 5.8 (1.92–24.63) | 629 (0.0) | NR | NR | NR | NR |
| Secondary mental or behavioural syndromes associated with disorders or diseases classified elsewhere (6E6) | 29 954 (0.5) | 121 788 | 9 675 | 79.4 | 17.3 (16.41–19.53) | 4 052 (0.1) | 19 534 | 1 466 | 75.0 | 33.3 (31.20–53.87) |

Abbreviation: NR, not reported. Diagnostic groups correspond to ICD-11 mental, behavioural, and neurodevelopmental disorder categories. Person-time was dynamically allocated according to time-varying psychiatric care setting (outside vs specialist psychiatric care). Crude mortality rates were calculated from deaths and person-years. Age-standardized mortality rates were estimated by direct standardization to the age distribution of the study population and are presented with 95% CIs. Cells with fewer than 6 observations were not reported.

**Table S2. Diagnosis-specific mortality rates by age group**

| | 5–64 years | | | | | ≥65 years | | | | |
|---|---|---|---|---|---|---|---|---|---|---|
| | No. (%) | Person-years | No. of deaths | Mortality rate per 1000 person-years | | No. (%) | Person-years | No. of deaths | Mortality rate per 1000 person-years | |
| | | | | Crude | Age-standardized | | | | Crude | Age-standardized |
| Mental, behavioural or neurodevelopmental disorders (6) | 1 144 753 (24.2) | 6 036 284 | 20 163 | 3.3 | 3.8 (3.8–3.9) | 386 628 (23.7) | 1 543 225 | 155 642 | 100.9 | 72.6 (72.2–73.0) |
| Neurodevelopmental disorders (6A0) | 220 354 (4.7) | 1 016 848 | 1 281 | 1.3 | 5.8 (5.4–6.2) | 5 900 (0.4) | 20 356 | 1 894 | 93.0 | 89.2 (85.2–93.4) |
| Schizophrenia or other primary psychotic disorders (6A2) | 28 596 (0.6) | 159 354 | 1 239 | 7.8 | 7.7 (7.0–9.0) | 12 038 (0.7) | 52 527 | 4 628 | 88.1 | 68.4 (66.3–70.6) |
| Catatonia (6A4) | NR | NR | NR | NR | NR | 38 (0.0) | 111 | 17 | 152.6 | 166.3 (89.3–292.0) |
| Bipolar or related disorders (6A6) | 26 205 (0.6) | 139 675 | 638 | 4.6 | 4.6 (4.2–8.6) | 2 693 (0.2) | 11 936 | 654 | 54.8 | 62.2 (57.5–67.3) |
| Depressive disorders (6A7) | 506 748 (10.7) | 2 495 429 | 7 102 | 2.8 | 2.9 (2.8–3.0) | 83 830 (5.1) | 382 092 | 21 025 | 55.0 | 48.8 (48.1–49.5) |
| Anxiety or fear-related disorders (6B0) | 520 853 (11.0) | 2 338 250 | 4 354 | 1.9 | 2.6 (2.5–2.7) | 44 311 (2.7) | 190 276 | 7 695 | 40.4 | 39.1 (38.3–40.0) |
| Obsessive-compulsive or related disorders (6B2) | 29 351 (0.6) | 126 244 | 187 | 1.5 | 2.6 (2.1–3.1) | 1 370 (0.1) | 5 869 | 262 | 44.6 | 46.5 (41.0–52.5) |
| Disorders specifically associated with stress (6B4) | 295 895 (6.2) | 1 252 207 | 3 563 | 2.8 | 2.6 (2.5–2.7) | 30 950 (1.9) | 131 678 | 4 781 | 36.3 | 40.5 (39.4–41.7) |

| | 5–64 years | | | | | ≥65 years | | | | |
|---|---|---|---|---|---|---|---|---|---|---|
| | **No. (%)** | **Person-years** | **No. of deaths** | **Mortality rate per 1000 person-years** | | **No. (%)** | **Person-years** | **No. of deaths** | **Mortality rate per 1000 person-years** | |
| | | | | **Crude** | **Age-standardized** | | | | **Crude** | **Age-standardized** |
| Dissociative disorders (6B6) | 6 769 (0.1) | 36 792 | 88 | 2.4 | 2.7 (2.1–4.4) | 864 (0.1) | 4 308 | 258 | 59.9 | 53.7 (47.3–60.8) |
| Feeding or eating disorders (6B8) | 25 341 (0.5) | 132 897 | 147 | 1.1 | 3.2 (2.4–4.3) | 513 (0.0) | 1 874 | 198 | 105.7 | 102.9 (89.1–118.4) |
| Elimination disorders (6C0) | 29 220 (0.6) | 183 700 | 43 | 0.2 | 4.4 (2.2–8.4) | 957 (0.1) | 4 884 | 367 | 75.1 | 47.1 (40.8–54.7) |
| Disorders of bodily distress or bodily experience (6C2) | 108 073 (2.3) | 622 313 | 755 | 1.2 | 1.2 (1.1–1.3) | 19 249 (1.2) | 90 126 | 2 215 | 24.6 | 30.2 (28.9–31.5) |
| Disorders due to substance use (6C4) | 144 102 (3.0) | 745 767 | 10 445 | 14.0 | 11.3 (11.1–12.5) | 50 752 (3.1) | 214 783 | 15 962 | 74.3 | 86.6 (85.1–88.0) |
| Disorders due to addictive behaviours (6C5) | 2 717 (0.1) | 11 424 | 37 | 3.2 | 3.8 (2.4–36.8) | 133 (0.0) | 592 | 13 | 22.0 | 29.7 (15.3–53.6) |
| Impulse control disorders (6C7) | 4 663 (0.1) | 21 780 | 57 | 2.6 | 4.2 (2.9–5.9) | 247 (0.0) | 1 019 | 62 | 60.9 | 63.2 (48.2–81.8) |
| Disruptive behaviour or dissocial disorders (6C9) | 117 739 (2.5) | 541 447 | 727 | 1.3 | 2.7 (2.4–2.9) | 5 228 (0.3) | 20 270 | 1 004 | 49.5 | 47.1 (44.2–50.1) |
| Paraphilic disorders (6D3) | NR | NR | NR | NR | NR | NR | NR | NR | NR | NR |
| Factitious disorders (6D5) | NR | NR | NR | NR | NR | 41 (0.0) | 198 | 15 | 75.8 | 69.5 (38.0–122.3) |
| Dementia (6D8) | 6 615 (0.1) | 20 471 | 1 042 | 50.9 | NR | 197 756 (12.1) | 646 200 | 112 953 | 174.8 | NR |

| | 5–64 years | | | | | ≥65 years | | | | |
|---|---|---|---|---|---|---|---|---|---|---|
| | **No. (%)** | **Person-years** | **No. of deaths** | **Mortality rate per 1000 person-years** | | **No. (%)** | **Person-years** | **No. of deaths** | **Mortality rate per 1000 person-years** | |
| | | | | **Crude** | **Age-standardized** | | | | **Crude** | **Age-standardized** |
| Mental or behavioural disorders associated with pregnancy, childbirth or the puerperium (6E2) | 1 640 (0.0) | 10 198 | 7 | 0.7 | 0.4 (0.1–18.3) | 21 (0.0) | 109 | 6 | 55.2 | 25.3 (8.3–114.7) |
| Secondary mental or behavioural syndromes associated with disorders or diseases classified elsewhere (6E6) | 5 673 (0.1) | 26 812 | 423 | 15.8 | 8.9 (8.0–11.6) | 28 976 (1.8) | 114 510 | 10 718 | 93.6 | 65.6 (63.8–67.5) |

Abbreviation: NR, not reported. Diagnostic groups correspond to ICD-11 mental, behavioural, and neurodevelopmental disorder categories. Crude mortality rates were calculated from deaths and person-years. Age-standardized mortality rates were estimated by direct standardization to the age distribution of the study population and are presented with 95% CIs. Cells with fewer than 6 observations were not reported. Age-standardization for dementia in the 5–64-year age group was unstable.

**Table S3. Diagnosis-specific mortality rates by gender**

| | Men | | | | | Women | | | | |
|---|---|---|---|---|---|---|---|---|---|---|
| | No. (%) | Person-years | No. of deaths | Mortality rate per 1000 person-years | | No. (%) | Person-years | No. of deaths | Mortality rate per 1000 person-years | |
| | | | | Crude | Age-standardized | | | | Crude | Age-standardized |
| Mental, behavioural or neurodevelopmental disorders (6) | 625 741 (22.8) | 3 205 442 | 84 650 | 26.4 | 22.1 (22.0–22.3) | 837 323 (30.1) | 4 374 068 | 91 155 | 20.8 | 15.6 (15.5–15.7) |
| Neurodevelopmental disorders (6A0) | 134 219 (4.9) | 668 567 | 1 781 | 2.7 | 24.1 (22.8–25.4) | 90 709 (3.3) | 368 637 | 1 394 | 3.8 | 22.2 (20.9–23.5) |
| Schizophrenia or other primary psychotic disorders (6A2) | 18 609 (0.7) | 103 241 | 2 438 | 23.6 | 25.8 (24.8–27.7) | 20 110 (0.7) | 108 640 | 3 429 | 31.6 | 18.0 (16.7–20.4) |
| Catatonia (6A4) | 40 (0.0) | 177 | 11 | 62.2 | 55.4 (24.4–299.0) | 28 (0.0) | 106 | 10 | 94.4 | 189.1 (21.2–797.0) |
| Bipolar or related disorders (6A6) | 11 630 (0.4) | 63 909 | 761 | 11.9 | 19.8 (18.1–25.3) | 16 120 (0.6) | 87 702 | 531 | 6.1 | 14.5 (13.1–22.3) |
| Depressive disorders (6A7) | 198 521 (7.2) | 997 187 | 12 085 | 12.1 | 16.0 (15.7–16.4) | 365 291 (13.1) | 1 880 335 | 16 042 | 8.5 | 11.5 (11.3–12.0) |
| Anxiety or fear-related disorders (6B0) | 203 770 (7.4) | 965 093 | 5 015 | 5.2 | 13.0 (12.6–13.4) | 348 053 (12.5) | 1 563 433 | 7 034 | 4.5 | 9.9 (9.6–10.1) |
| Obsessive-compulsive or related disorders (6B2) | 11 028 (0.4) | 51 359 | 232 | 4.5 | 12.2 (10.5–14.2) | 19 203 (0.7) | 80 754 | 217 | 2.7 | 11.7 (10.0–13.6) |
| Disorders specifically associated with stress (6B4) | 95 672 (3.5) | 405 598 | 3 469 | 8.6 | 14.8 (14.2–15.3) | 217 253 (7.8) | 978 287 | 4 875 | 5.0 | 9.8 (9.4–10.1) |
| Dissociative disorders (6B6) | 2 026 (0.1) | 11 599 | 139 | 12.0 | 15.2 (12.7–19.1) | 5 323 (0.2) | 29 501 | 207 | 7.0 | 13.1 (11.2–16.6) |

| | Men | | | | | Women | | | | |
|---|---|---|---|---|---|---|---|---|---|---|
| | No. (%) | Person-years | No. of deaths | Mortality rate per 1000 person-years | | No. (%) | Person-years | No. of deaths | Mortality rate per 1000 person-years | |
| | | | | Crude | Age-standardized | | | | Crude | Age-standardized |
| Feeding or eating disorders (6B8) | 2 863 (0.1) | 14 636 | 124 | 8.5 | 36.0 (29.2–44.0) | 22 868 (0.8) | 120 136 | 221 | 1.8 | 23.0 (19.2–27.4) |
| Elimination disorders (6C0) | 18 510 (0.7) | 116 227 | 152 | 1.3 | 21.8 (15.8–31.2) | 11 601 (0.4) | 72 357 | 258 | 3.6 | 11.1 (8.9–15.2) |
| Disorders of bodily distress or bodily experience (6C2) | 34 493 (1.3) | 204 097 | 1 185 | 5.8 | 8.1 (7.6–8.6) | 85 849 (3.1) | 508 343 | 1 785 | 3.5 | 7.5 (7.1–7.9) |
| Disorders due to substance use (6C4) | 119 024 (4.3) | 644 325 | 20 271 | 31.5 | 27.8 (27.4–29.5) | 58 859 (2.1) | 316 225 | 6 136 | 19.4 | 21.3 (20.7–23.4) |
| Disorders due to addictive behaviours (6C5) | 2 095 (0.1) | 8 839 | 38 | 4.3 | 9.3 (4.8–47.3) | 698 (0.0) | 3 177 | 12 | 3.8 | 8.5 (4.0–176.9) |
| Impulse control disorders (6C7) | 3 352 (0.1) | 15 373 | 84 | 5.5 | 21.2 (15.9–28.0) | 1 478 (0.1) | 7 425 | 35 | 4.7 | 11.6 (7.7–16.9) |
| Disruptive behaviour or dissocial disorders (6C9) | 50 407 (1.8) | 257 923 | 826 | 3.2 | 17.0 (15.6–18.4) | 71 162 (2.6) | 303 794 | 905 | 3.0 | 10.5 (9.8–11.3) |
| Personality disorders and related traits (6D10-6D11) | 11 927 (0.4) | 65 988 | 512 | 7.8 | 20.2 (17.6–25.1) | 24 168 (0.9) | 116 908 | 432 | 3.7 | 18.6 (16.1–27.7) |
| Paraphilic disorders (6D3) | 88 (0.0) | 472 | 6 | 12.7 | 30.6 (9.9–1 316.0) | 23 (0.0) | NR | NR | NR | NR |
| Factitious disorders (6D5) | 340 (0.0) | 970 | 8 | 8.2 | 22.2 (7.5–93.9) | 429 (0.0) | 1 250 | 8 | 6.4 | 15.7 (6.5–91.8) |

| | Men | | | | | Women | | | | |
|---|---|---|---|---|---|---|---|---|---|---|
| | **No. (%)** | **Person-years** | **No. of deaths** | **Mortality rate per 1000 person-years** | | **No. (%)** | **Person-years** | **No. of deaths** | **Mortality rate per 1000 person-years** | |
| | | | | **Crude** | **Age-standardized** | | | | **Crude** | **Age-standardized** |
| Neurocognitive disorders (6D7-6E0) | 99 049 (3.6) | 326 371 | 56 097 | 171.9 | 40.1 (37.9–44.1) | 137 928 (5.0) | 499 853 | 70 346 | 140.7 | 30.8 (29.6–35.1) |
| Mental or behavioural disorders associated with pregnancy, childbirth or the puerperium (6E2) | 31 (0.0) | NR | NR | NR | NR | 1 625 (0.1) | 10 103 | 11 | 1.1 | 5.9 (1.7–35.3) |
| Secondary mental or behavioural syndromes associated with disorders or diseases classified elsewhere (6E6) | 14 865 (0.5) | 61 457 | 5 401 | 87.9 | 22.8 (21.5–27.5) | 18 327 (0.7) | 79 866 | 5 740 | 71.9 | 18.1 (17.1–21.6) |
| Mental, behavioural or neurodevelopmental disorders (6)—specialist psychiatric care only | 170 593 (6.2) | 1 007 482 | 11 921 | 11.8 | 24.0 (23.5–24.4) | 234 818 (8.4) | 1 394 063 | 12 148 | 8.7 | 16.9 (16.6–17.2) |

Abbreviation: NR, not reported. Diagnostic groups correspond to ICD-11 mental, behavioural, and neurodevelopmental disorder categories. Crude mortality rates were calculated from deaths and person-years. Age-standardized mortality rates were estimated by direct standardization to the age distribution of the study population and are presented with 95% CIs. Cells with fewer than 6 observations were not reported. The final row is restricted to diagnoses recorded in specialist psychiatric care.

**Table S4. Diagnosis-specific mortality rates by substance use disorder status**

| | No SUD | | | | | SUD | | | | |
|---|---|---|---|---|---|---|---|---|---|---|
| | No. (%) | Person-years | No. of deaths | Mortality rate per 1000 person-years | | No. (%) | Person-years | No. of deaths | Mortality rate per 1000 person-years | |
| | | | | Crude | Age-standardized | | | | Crude | Age-standardized |
| Mental, behavioural or neurodevelopmental disorders (6) | 1 338 717 (24.2) | 6 618 959 | 149 398 | 22.6 | 16.0 (15.89–16.08) | 177 883 (3.2) | 960 550 | 26 407 | 27.5 | 27.0 (26.62–27.97) |
| Neurodevelopmental disorders (6A0) | 218 153 (3.9) | 996 060 | 2 819 | 2.8 | 22.6 (21.71–23.56) | 12 949 (0.2) | 41 144 | 356 | 8.7 | 32.9 (27.85–40.11) |
| Schizophrenia or other primary psychotic disorders (6A2) | 32 898 (0.6) | 169 817 | 4 976 | 29.3 | 18.2 (17.44–19.32) | 8 768 (0.2) | 42 065 | 891 | 21.2 | 33.3 (30.70–106.27) |
| Catatonia (6A4) | 65 (0.0) | 255 | 21 | 82.4 | 53.9 (30.97–287.68) | 9 (0.0) | NR | NR | NR | NR |
| Bipolar or related disorders (6A6) | 24 016 (0.4) | 122 216 | 895 | 7.3 | 14.9 (13.87–18.25) | 6 292 (0.1) | 29 395 | 397 | 13.5 | 25.6 (21.69–267.86) |
| Depressive disorders (6A7) | 536 477 (9.7) | 2 599 947 | 23 284 | 9.0 | 11.2 (11.03–11.37) | 59 397 (1.1) | 277 575 | 4 843 | 17.4 | 25.0 (24.21–42.80) |
| Anxiety or fear-related disorders (6B0) | 527 610 (9.5) | 2 318 628 | 9 451 | 4.1 | 9.1 (8.94–9.33) | 49 475 (0.9) | 209 897 | 2 598 | 12.4 | 21.5 (20.38–26.31) |
| Obsessive-compulsive or related disorders (6B2) | 29 007 (0.5) | 123 606 | 368 | 3.0 | 11.0 (9.82–12.34) | 2 225 (0.0) | 8 507 | 81 | 9.5 | 24.5 (15.74–41.33) |
| Disorders specifically associated with stress (6B4) | 300 327 (5.4) | 1 293 194 | 7 145 | 5.5 | 9.9 (9.61–10.13) | 23 605 (0.4) | 90 691 | 1 199 | 13.2 | 25.2 (23.12–27.69) |
| Dissociative disorders (6B6) | 6 884 (0.1) | 37 172 | 306 | 8.2 | 12.6 (11.22–14.75) | 895 (0.0) | 3 928 | 40 | 10.2 | 29.0 (17.34–49.33) |

| | No SUD | | | | | SUD | | | | |
|---|---|---|---|---|---|---|---|---|---|---|
| | No. (%) | Person-years | No. of deaths | Mortality rate per 1000 person-years | | No. (%) | Person-years | No. of deaths | Mortality rate per 1000 person-years | |
| | | | | Crude | Age-standardized | | | | Crude | Age-standardized |
| Feeding or eating disorders (6B8) | 24 941 (0.5) | 126 178 | 290 | 2.3 | 22.8 (19.84–26.16) | 2 086 (0.0) | 8 593 | 55 | 6.4 | 43.2 (23.54–733.51) |
| Elimination disorders (6C0) | 30 075 (0.5) | 187 293 | 392 | 2.1 | 12.9 (10.52–16.33) | 469 (0.0) | 1 291 | 18 | 13.9 | 31.3 (15.14–157.83) |
| Disorders of bodily distress or bodily experience (6C2) | 117 790 (2.1) | 692 193 | 2 644 | 3.8 | 6.8 (6.55–7.14) | 5 129 (0.1) | 20 247 | 326 | 16.1 | 21.7 (19.11–88.21) |
| Disorders due to substance use (6C4) | | | | | | 177 883 (3.2) | 960 550 | 26 407 | 27.5 | 27.0 (26.62–27.97) |
| Disorders due to addictive behaviours (6C5) | 2 286 (0.0) | 9 109 | 35 | 3.8 | 8.6 (5.27–45.08) | 696 (0.0) | 2 906 | 15 | 5.2 | 9.6 (2.46–115.56) |
| Impulse control disorders (6C7) | 4 210 (0.1) | 18 764 | 91 | 4.8 | 15.8 (12.34–20.05) | 939 (0.0) | 4 034 | 28 | 6.9 | 15.2 (7.39–112.49) |
| Disruptive behaviour or dissocial disorders (6C9) | 116 145 (2.1) | 514 605 | 1 295 | 2.5 | 10.7 (10.03–11.31) | 12 065 (0.2) | 47 112 | 436 | 9.3 | 24.8 (21.59–44.12) |
| Personality disorders and related traits (6D10-6D11) | 30 551 (0.6) | 145 081 | 522 | 3.6 | 16.7 (14.83–20.26) | 7 923 (0.1) | 37 815 | 422 | 11.2 | 29.2 (24.27–122.23) |
| Paraphilic disorders (6D3) | 94 (0.0) | NR | NR | NR | NR | 18 (0.0) | NR | NR | NR | NR |
| Factitious disorders (6D5) | 697 (0.0) | 1 885 | 13 | 6.9 | 13.4 (6.88–49.30) | 100 (0.0) | NR | NR | NR | NR |
| Neurocognitive disorders (6D7-6E0) | 227 572 (4.1) | 781 626 | 121 202 | 155.1 | 31.7 (30.37–34.06) | 12 591 (0.2) | 44 598 | 5 241 | 117.5 | 55.7 (52.98–140.77) |

| | No SUD | | | | | SUD | | | | |
|---|---|---|---|---|---|---|---|---|---|---|
| | **No. (%)** | **Person-years** | **No. of deaths** | **Mortality rate per 1000 person-years** | | **No. (%)** | **Person-years** | **No. of deaths** | **Mortality rate per 1000 person-years** | |
| | | | | **Crude** | **Age-standardized** | | | | **Crude** | **Age-standardized** |
| Mental or behavioural disorders associated with pregnancy, childbirth or the puerperium (6E2) | 1 621 (0.0) | 10 024 | 12 | 1.2 | 5.5 (1.92–24.70) | 76 (0.0) | NR | NR | NR | NR |
| Secondary mental or behavioural syndromes associated with disorders or diseases classified elsewhere (6E6) | 31 580 (0.6) | 132 143 | 10 473 | 79.3 | 18.9 (18.06–20.99) | 2 325 (0.0) | 9 180 | 668 | 72.8 | 37.2 (33.60–189.94) |

Abbreviation: NR, not reported. Diagnostic groups correspond to ICD-11 mental, behavioural, and neurodevelopmental disorder categories. Person-time was dynamically allocated according to time-varying substance use disorder (SUD) status. Crude mortality rates were calculated from deaths and person-years. Age-standardized mortality rates were estimated by direct standardization to the age distribution of the study population and are presented with 95% CIs. Cells with fewer than 6 observations were not reported.

**Table S5. Mortality rates by regression model covariates**

| | No. (%) | Person-years | No. of deaths | Mortality rate per 1000 person-years | |
|---|---|---|---|---|---|
| | | | | Crude | Age-standardized |
| **Gender** | | | | | |
| Men | 2 741 702 (49.6) | 31 162 920 | 260 621 | 8.36 | 10.27 (10.23–10.31) |
| Women | 2 784 897 (50.4) | 31 852 225 | 237 947 | 7.47 | 6.20 (6.18–6.23) |
| **Region** | | | | | |
| 0 | 411 027 (7.4) | 4 275 801 | 31 580 | 7.4 | 54.5 (53.84–55.12) |
| 1 | 1 960 763 (35.5) | 22 456 483 | 162 296 | 7.2 | 7.5 (7.48–7.56) |
| 2 | 849 283 (15.4) | 9 739 405 | 82 972 | 8.5 | 7.3 (7.25–7.35) |
| 3 | 834 265 (15.1) | 9 606 303 | 78 266 | 8.1 | 7.4 (7.32–7.42) |
| 4 | 769 409 (13.9) | 8 873 114 | 81 403 | 9.2 | 7.8 (7.71–7.82) |
| 5 | 701 852 (12.7) | 8 064 039 | 62 051 | 7.7 | 7.6 (7.53–7.65) |
| **Charlson Comorbidity Index** | | | | | |
| 0 | 4 100 679 (74.2) | 47 531 951 | 73 615 | 1.5 | 3.5 (3.43–3.49) |
| 1–3 | 1 171 249 (21.2) | 13 113 494 | 262 679 | 20.0 | 9.0 (8.91–8.99) |
| >= 4 | 254 671 (4.6) | 2 369 699 | 162 274 | 68.5 | 29.5 (28.91–30.09) |
| **Period** | | | | | |
| 2011–2014 | 4 988 993 | 19 133 729 | 127 066 | 6.6 | 7.6 (7.56–7.64) |
| 2015–2019 | 5 148 255 | 24 335 781 | 191 627 | 7.9 | 7.8 (7.80–7.87) |
| 2020–2023 | 5 110 450 | 19 545 634 | 179 875 | 9.2 | 8.2 (8.17–8.24) |
| **Age-group** | | | | | |
| 5–9 | | 4 046 956 | 262 | 0.06 | |
| 10–14 | | 3 980 666 | 305 | 0.08 | |
| 15–19 | | 3 982 820 | 1091 | 0.27 | |
| 20–24 | | 4 194 239 | 1646 | 0.39 | |
| 25–29 | | 4 387 431 | 1611 | 0.37 | |
| 30–34 | | 4 375 917 | 1742 | 0.40 | |
| 35–39 | | 4 225 207 | 2227 | 0.53 | |
| 40–44 | | 4 017 027 | 3277 | 0.82 | |
| 45–49 | | 4 039 143 | 5405 | 1.34 | |

| | No. (%) | Person-years | No. of deaths | Mortality rate per 1000 person-years | |
|---|---|---|---|---|---|
| | | | | Crude | Age-standardized |
| 50–54 | | 4 180 485 | 9348 | 2.24 | |
| 55–59 | | 4 242 006 | 15646 | 3.69 | |
| 60–64 | | 4 209 065 | 25813 | 6.13 | |
| 65–69 | | 4 011 596 | 39466 | 9.84 | |
| 70–74 | | 3 362 104 | 53217 | 15.83 | |
| 75–79 | | 2 484 489 | 67155 | 27.03 | |
| 80–84 | | 1 773 774 | 88786 | 50.05 | |
| 85–89 | | 1 065 322 | 102428 | 96.15 | |
| 90–94 | | 436 896 | 79143 | 181.15 | |

No. (%) is shown for individual-level covariates (gender, region, and Charlson Comorbidity Index [CCI]); person-years are shown for time-varying covariates (calendar period and age group). Crude mortality rates were calculated from deaths and person-years. Age-standardized mortality rates were estimated by direct standardization to the age distribution of the study population and are presented with 95% CIs. Regions were grouped as follows: 1, Southern Finland; 2, Western Finland; 3, Central Finland; 4, Eastern Finland; and 5, Northern Finland. Region 0 included individuals registered in the Finnish Population Information System who did not belong to the resident population, such as those living in institutions or without a municipality of residence. CCI categories were 0, 1 to 3, and 4 or greater; 0 indicates no Charlson comorbidities.

**Table S6. Diagnosis-specific mortality rate ratios by psychiatric care setting and substance use disorder status among individuals aged 5 to 64 years**

| | Mortality rate ratio (95% CI) | | | | | | | |
|---|---|---|---|---|---|---|---|---|
| | Men | | | | Women | | | |
| | Outside specialist psychiatric care | | Specialist psychiatric care | | Outside specialist psychiatric care | | Specialist psychiatric care | |
| | No SUD | SUD | No SUD | SUD | No SUD | SUD | No SUD | SUD |
| Mental, behavioural or neurodevelopmental disorders (6) | 1.93 (1.76–2.12) | 6.34 (5.61–7.17) | 3.07 (2.77–3.41) | 8.97 (7.86–10.24) | 1.54 (1.39–1.71) | 5.15 (4.28–6.20) | 2.75 (2.46–3.08) | 7.29 (6.09–8.73) |
| Neurodevelopmental disorders (6A0) | 3.15 (2.75–3.62) | 5.54 (4.27–7.19) | 1.48 (1.19–1.85) | 9.49 (7.63–11.79) | 4.79 (4.11–5.60) | 5.75 (3.56–9.27) | 1.51 (1.08–2.10) | 6.61 (4.00–10.94) |
| Schizophrenia or other primary psychotic disorders (6A2) | 3.64 (2.87–4.61) | 12.10 (9.48–15.45) | 4.45 (3.81–5.19) | 12.34 (10.38–14.67) | 2.96 (2.05–4.27) | 13.37 (8.36–21.38) | 3.66 (3.03–4.43) | 10.88 (8.18–14.45) |
| Bipolar or related disorders (6A6) | 2.35 (1.77–3.10) | 6.18 (4.60–8.30) | 2.57 (2.12–3.12) | 5.93 (4.74–7.43) | 1.89 (1.36–2.62) | 7.19 (4.17–12.41) | 2.07 (1.63–2.63) | 8.25 (5.94–11.46) |
| Depressive disorders (6A7) | 1.30 (1.17–1.43) | 5.84 (5.14–6.64) | 1.89 (1.67–2.14) | 6.08 (5.24–7.06) | 1.06 (0.96–1.18) | 4.72 (3.86–5.78) | 1.69 (1.51–1.88) | 5.62 (4.62–6.83) |
| Anxiety or fear-related disorders (6B0) | 1.27 (1.14–1.41) | 6.28 (5.45–7.23) | 1.90 (1.68–2.14) | 7.65 (6.56–8.93) | 1.21 (1.08–1.36) | 5.45 (4.48–6.63) | 1.99 (1.75–2.27) | 7.26 (5.83–9.03) |
| Obsessive-compulsive or related disorders (6B2) | 1.16 (0.77–1.73) | 7.92 (5.22–12.02) | 1.62 (1.19–2.20) | 5.95 (3.96–8.93) | 1.43 (0.96–2.13) | 4.02 (1.36–11.87) | 1.72 (1.22–2.43) | 8.81 (4.75–16.35) |
| Disorders specifically associated with stress (6B4) | 1.06 (0.94–1.20) | 4.96 (4.24–5.81) | 3.01 (2.64–3.44) | 5.70 (4.76–6.83) | 1.01 (0.89–1.13) | 4.31 (3.36–5.53) | 3.37 (2.95–3.85) | 7.17 (5.77–8.91) |
| Dissociative disorders (6B6) | 0.87 (0.43–1.78) | 3.70 (1.55–8.83) | 1.61 (0.91–2.86) | 4.85 (2.35–10.01) | 1.33 (0.82–2.15) | 16.42 (7.35–36.70) | 1.71 (1.08–2.69) | 7.91 (3.77–16.62) |

| | Mortality rate ratio (95% CI) | | | | | | | |
|---|---|---|---|---|---|---|---|---|
| | **Men** | | | | **Women** | | | |
| | **Outside specialist psychiatric care** | | **Specialist psychiatric care** | | **Outside specialist psychiatric care** | | **Specialist psychiatric care** | |
| | **No SUD** | **SUD** | **No SUD** | **SUD** | **No SUD** | **SUD** | **No SUD** | **SUD** |
| Feeding or eating disorders (6B8) | 2.93 (1.72–5.00) | 7.24 (2.38–22.03) | 2.48 (1.51–4.06) | 13.38 (6.33–28.30) | 2.51 (1.70–3.69) | 4.93 (1.91–12.75) | 1.99 (1.45–2.74) | 9.92 (6.35–15.50) |
| Disorders of bodily distress or bodily experience (6C2) | 0.62 (0.53–0.73) | 4.50 (3.60–5.63) | 1.61 (1.08–2.41) | 3.63 (1.85–7.14) | 0.74 (0.65–0.85) | 3.41 (2.40–4.84) | 1.34 (0.85–2.13) | 9.03 (4.60–17.72) |
| Disorders due to substance use (6C4) | NA | 5.73 (5.10–6.44) | NA | 8.66 (7.61–9.85) | NA | 4.47 (3.78–5.28) | NA | 7.03 (5.83–8.47) |
| Disorders due to addictive behaviours (6C5) | 1.96 (1.00–3.83) | 3.19 (1.24–8.16) | 3.48 (2.02–6.02) | 4.12 (2.02–8.40) | 1.11 (0.29–4.24) | NR | 1.37 (0.34–5.45) | NR |
| Impulse control disorders (6C7) | 1.55 (0.85–2.82) | 5.19 (2.50–10.79) | 3.23 (1.99–5.24) | 3.83 (1.96–7.48) | 2.36 (1.07–5.22) | 7.08 (1.69–29.65) | 0.80 (0.20–3.23) | 1.83 (0.25–13.33) |
| Disruptive behaviour or dissocial disorders (6C9) | 1.39 (1.12–1.73) | 6.43 (5.08–8.12) | 1.62 (1.31–2.02) | 8.27 (6.53–10.47) | 1.30 (1.02–1.64) | 6.92 (4.66–10.29) | 2.26 (1.79–2.84) | 10.91 (7.63–15.61) |
| Personality disorders and related traits (6D10-6D11) | 1.32 (0.98–1.77) | 6.84 (5.40–8.67) | 1.57 (1.28–1.93) | 6.22 (5.06–7.65) | 1.42 (0.99–2.04) | 6.61 (4.26–10.26) | 1.70 (1.33–2.17) | 7.49 (5.56–10.07) |
| Neurocognitive disorders (6D7-6E0) | 4.27 (3.69–4.93) | 8.05 (6.59–9.83) | 6.23 (4.84–8.03) | 9.39 (7.24–12.19) | 5.72 (4.66–7.02) | 8.65 (6.24–11.99) | 7.93 (5.78–10.87) | 6.09 (3.54–10.46) |
| Dementia (6D8) | 5.52 (4.59–6.63) | 6.52 (5.03–8.44) | 7.41 (5.08–10.81) | 6.55 (3.82–11.24) | 8.11 (6.44–10.21) | 5.45 (3.38–8.79) | 10.44 (6.51–16.73) | 2.37 (0.58–9.70) |

| | Mortality rate ratio (95% CI) | | | | | | | |
|---|---|---|---|---|---|---|---|---|
| | **Men** | | | | **Women** | | | |
| | **Outside specialist psychiatric care** | | **Specialist psychiatric care** | | **Outside specialist psychiatric care** | | **Specialist psychiatric care** | |
| | **No SUD** | **SUD** | **No SUD** | **SUD** | **No SUD** | **SUD** | **No SUD** | **SUD** |
| Secondary mental or behavioural syndromes associated with disorders or diseases classified elsewhere (6E6) | 2.43 (1.97–3.01) | 4.33 (3.09–6.07) | 3.64 (2.85–4.65) | 5.99 (4.47–8.02) | 2.24 (1.73–2.91) | 4.52 (2.43–8.40) | 5.59 (4.16–7.52) | 10.97 (6.47–18.60) |

Abbreviations: MRR, mortality rate ratio; NA, not applicable; NR, not reported; SUD, substance use disorder.

Diagnostic groups correspond to ICD-11 mental, behavioural, and neurodevelopmental disorder categories. Person-time was dynamically allocated according to time-varying psychiatric care setting and SUD status. MRRs were estimated separately for women and men using Poisson regression adjusted for age group, calendar period, region, and CCI. For disorders due to substance use (6C4), estimates without SUD were not applicable. Diagnostic groups with insufficient deaths for stable estimation were not reported

**Table S7. Diagnosis-specific mortality rate ratios by psychiatric care setting and substance use disorder status among individuals aged 65 to 95 years**

| | Men | | | | Women | | | |
|---|---|---|---|---|---|---|---|---|
| | Outside specialist psychiatric care | | Specialist psychiatric care | | Outside specialist psychiatric care | | Specialist psychiatric care | |
| | No SUD | SUD | No SUD | SUD | No SUD | SUD | No SUD | SUD |
| Mental, behavioural or neurodevelopmental disorders (6) | 2.67 (2.50–2.85) | 3.10 (2.86–3.36) | 2.98 (2.76–3.20) | 3.43 (3.09–3.80) | 2.37 (2.21–2.55) | 2.90 (2.63–3.20) | 2.42 (2.23–2.63) | 3.00 (2.63–3.44) |
| Neurodevelopmental disorders (6A0) | 2.40 (2.14–2.69) | 3.15 (2.47–4.02) | 2.75 (1.78–4.26) | 2.46 (0.96–6.33) | 2.07 (1.79–2.39) | 2.64 (1.87–3.74) | 2.85 (1.60–5.09) | NR |
| Schizophrenia or other primary psychotic disorders (6A2) | 2.23 (2.01–2.47) | 3.02 (2.32–3.94) | 2.15 (1.93–2.39) | 2.52 (1.95–3.25) | 1.66 (1.50–1.85) | 1.79 (1.27–2.52) | 1.66 (1.51–1.83) | 2.69 (2.10–3.43) |
| Bipolar or related disorders (6A6) | 2.08 (1.75–2.47) | 2.78 (2.10–3.68) | 1.69 (1.39–2.04) | 1.91 (1.35–2.72) | 1.62 (1.36–1.94) | 2.50 (1.59–3.92) | 1.70 (1.39–2.06) | 1.12 (0.45–2.81) |
| Depressive disorders (6A7) | 1.41 (1.32–1.50) | 2.29 (2.07–2.54) | 1.70 (1.57–1.83) | 2.41 (2.10–2.76) | 1.27 (1.18–1.35) | 2.04 (1.76–2.37) | 1.50 (1.39–1.63) | 2.13 (1.81–2.50) |
| Anxiety or fear-related disorders (6B0) | 1.19 (1.10–1.29) | 2.00 (1.71–2.34) | 1.31 (1.17–1.47) | 1.95 (1.59–2.38) | 1.06 (0.98–1.14) | 1.86 (1.58–2.20) | 1.36 (1.22–1.51) | 1.57 (1.22–2.02) |
| Obsessive-compulsive or related disorders (6B2) | 1.14 (0.91–1.44) | 2.57 (1.52–4.36) | 1.42 (0.99–2.05) | 0.59 (0.17–2.04) | 1.47 (1.22–1.77) | 1.28 (0.41–4.04) | 1.52 (1.08–2.14) | 0.51 (0.06–4.02) |
| Disorders specifically associated with stress (6B4) | 1.04 (0.95–1.14) | 2.21 (1.80–2.71) | 2.18 (1.93–2.46) | 2.71 (2.17–3.38) | 0.86 (0.78–0.95) | 2.02 (1.62–2.52) | 2.09 (1.84–2.38) | 2.50 (1.89–3.30) |
| Dissociative disorders (6B6) | 1.54 (1.25–1.89) | 1.88 (0.82–4.29) | 1.90 (0.88–4.12) | 2.99 (0.38–23.44) | 1.34 (1.12–1.61) | 2.22 (0.89–5.56) | 1.76 (1.12–2.78) | NR |
| Feeding or eating disorders (6B8) | 3.40 (2.52–4.58) | 8.70 (4.99–15.14) | 0.88 (0.11–6.86) | NR | 2.63 (2.11–3.27) | 2.71 (1.03–7.13) | 1.66 (0.78–3.52) | NR |

| | Men | | | | Women | | | |
|---|---|---|---|---|---|---|---|---|
| | Outside specialist psychiatric care | | Specialist psychiatric care | | Outside specialist psychiatric care | | Specialist psychiatric care | |
| | No SUD | SUD | No SUD | SUD | No SUD | SUD | No SUD | SUD |
| Disorders of bodily distress or bodily experience (6C2) | 0.83 (0.74–0.93) | 1.83 (1.44–2.32) | 1.34 (0.96–1.87) | 1.62 (0.67–3.95) | 0.83 (0.76–0.91) | 1.95 (1.51–2.51) | 1.76 (1.35–2.29) | 2.13 (0.91–4.95) |
| Disorders due to substance use (6C4) | NA | 2.55 (2.35–2.76) | NA | 2.58 (2.28–2.91) | NA | 2.23 (2.02–2.47) | NA | 2.32 (1.96–2.76) |
| Disorders due to addictive behaviours (6C5) | 1.32 (0.50–3.45) | NR | 1.06 (0.15–7.67) | NR | 0.69 (0.34–1.40) | 1.32 (0.16–11.22) | 0.46 (0.08–2.78) | 19.57 (4.35–88.02) |
| Impulse control disorders (6C7) | 2.69 (1.94–3.75) | 1.15 (0.27–4.87) | 1.72 (0.64–4.65) | NR | 0.88 (0.49–1.58) | 2.39 (0.72–7.94) | 1.31 (0.57–3.00) | 19.57 (4.35–88.02) |
| Disruptive behaviour or dissocial disorders (6C9) | 1.68 (1.45–1.94) | 2.63 (2.02–3.41) | 1.28 (0.97–1.69) | 1.83 (1.03–3.25) | 1.15 (1.01–1.31) | 2.30 (1.72–3.06) | 1.43 (1.15–1.77) | 0.57 (0.20–1.59) |
| Personality disorders and related traits (6D10-6D11) | 1.91 (1.48–2.45) | 3.07 (2.29–4.11) | 1.99 (1.48–2.67) | 1.91 (1.10–3.30) | 2.44 (1.99–2.99) | 2.58 (1.71–3.90) | 1.75 (1.29–2.38) | 3.35 (2.17–5.17) |
| Neurocognitive disorders (6D7-6E0) | 2.86 (2.67–3.06) | 3.55 (3.25–3.89) | 4.32 (3.92–4.75) | 4.29 (3.72–4.95) | 2.71 (2.51–2.93) | 3.13 (2.81–3.49) | 3.13 (2.80–3.50) | 3.24 (2.62–3.99) |
| Dementia (6D8) | 2.92 (2.72–3.13) | 3.43 (3.13–3.75) | 4.24 (3.83–4.69) | 4.47 (3.80–5.25) | 2.77 (2.56–3.00) | 3.08 (2.74–3.45) | 3.05 (2.72–3.41) | 3.00 (2.36–3.82) |
| Secondary mental or behavioural syndromes associated with disorders or diseases classified elsewhere (6E6) | 1.34 (1.25–1.44) | 2.05 (1.76–2.38) | 2.68 (2.33–3.09) | 3.09 (2.35–4.05) | 1.25 (1.16–1.34) | 1.88 (1.57–2.25) | 2.17 (1.87–2.50) | 1.44 (0.97–2.14) |

Abbreviations: MRR, mortality rate ratio; NA, not applicable; NR, not reported; SUD, substance use disorder.

Diagnostic groups correspond to ICD-11 mental, behavioural, and neurodevelopmental disorder categories. Person-time was dynamically allocated according to time-varying psychiatric care setting and SUD status. MRRs were estimated separately for women and men using Poisson regression adjusted for age group, calendar period, region, and Charlson Comorbidity Index. For disorders due to substance use (6C4), estimates without SUD were not applicable. Diagnostic groups with insufficient deaths for stable estimation were not reported

**Figure S1. Time-varying mortality rate ratios for neurodevelopmental disorders (6A0) by psychiatric care setting, substance use disorder status, and cause of death**

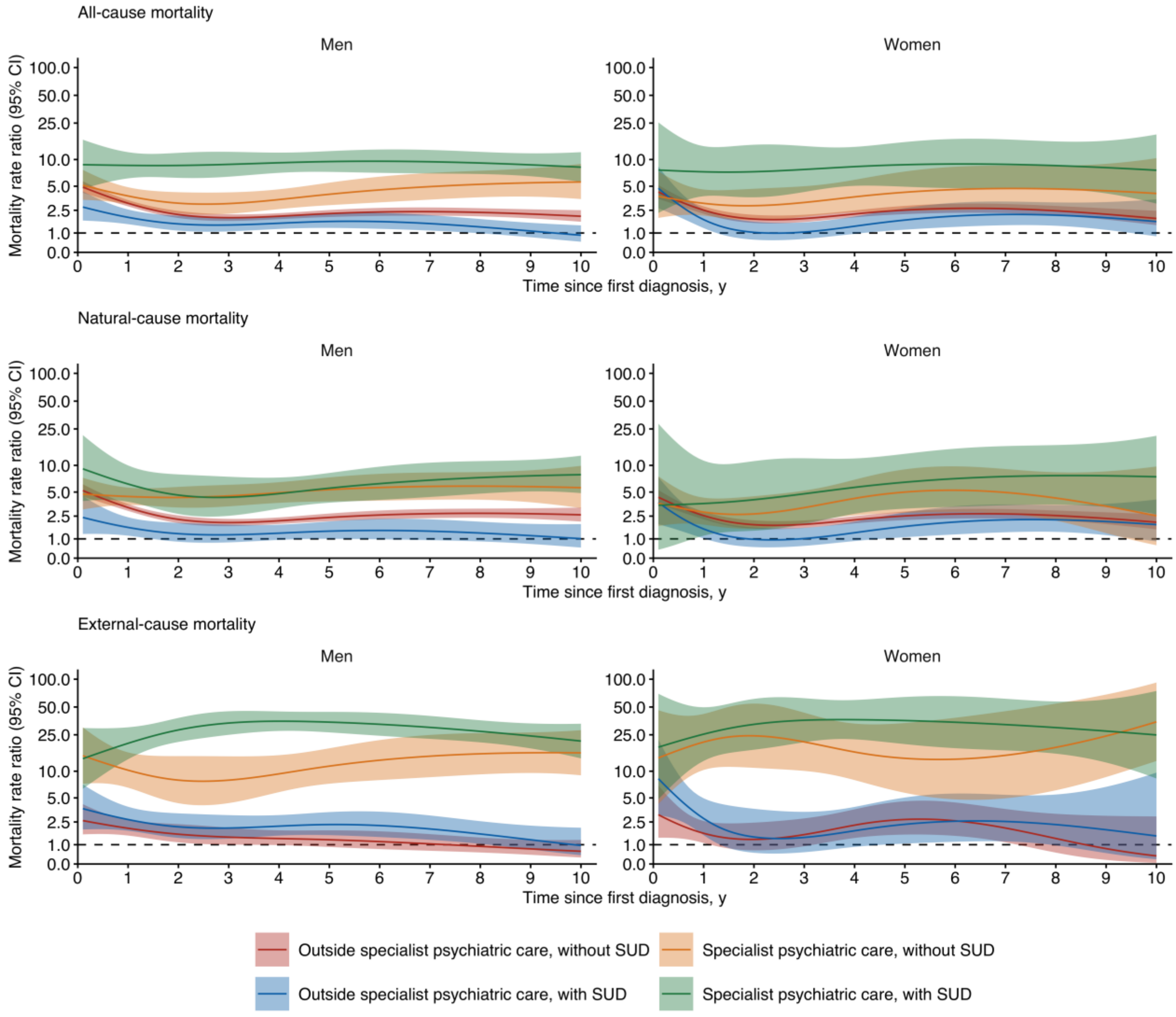

Abbreviations: MRR, mortality rate ratio; SUD, substance use disorder. MRRs are relative to individuals without the specific diagnostic group. Person-time was dynamically allocated according to time-varying psychiatric care setting and SUD status. MRRs were estimated separately for women and men using Poisson regression adjusted for age group, calendar period, region, and Charlson Comorbidity Index. Estimates are shown for the first 10 years after diagnosis. Shaded areas indicate 95% CIs; the dashed horizontal line indicates an MRR of 1. The y-axis is logarithmic.

**Figure S2. Time-varying mortality rate ratios for schizophrenia or other primary psychotic disorders (6A2) by psychiatric care setting, substance use disorder status, and cause of death**

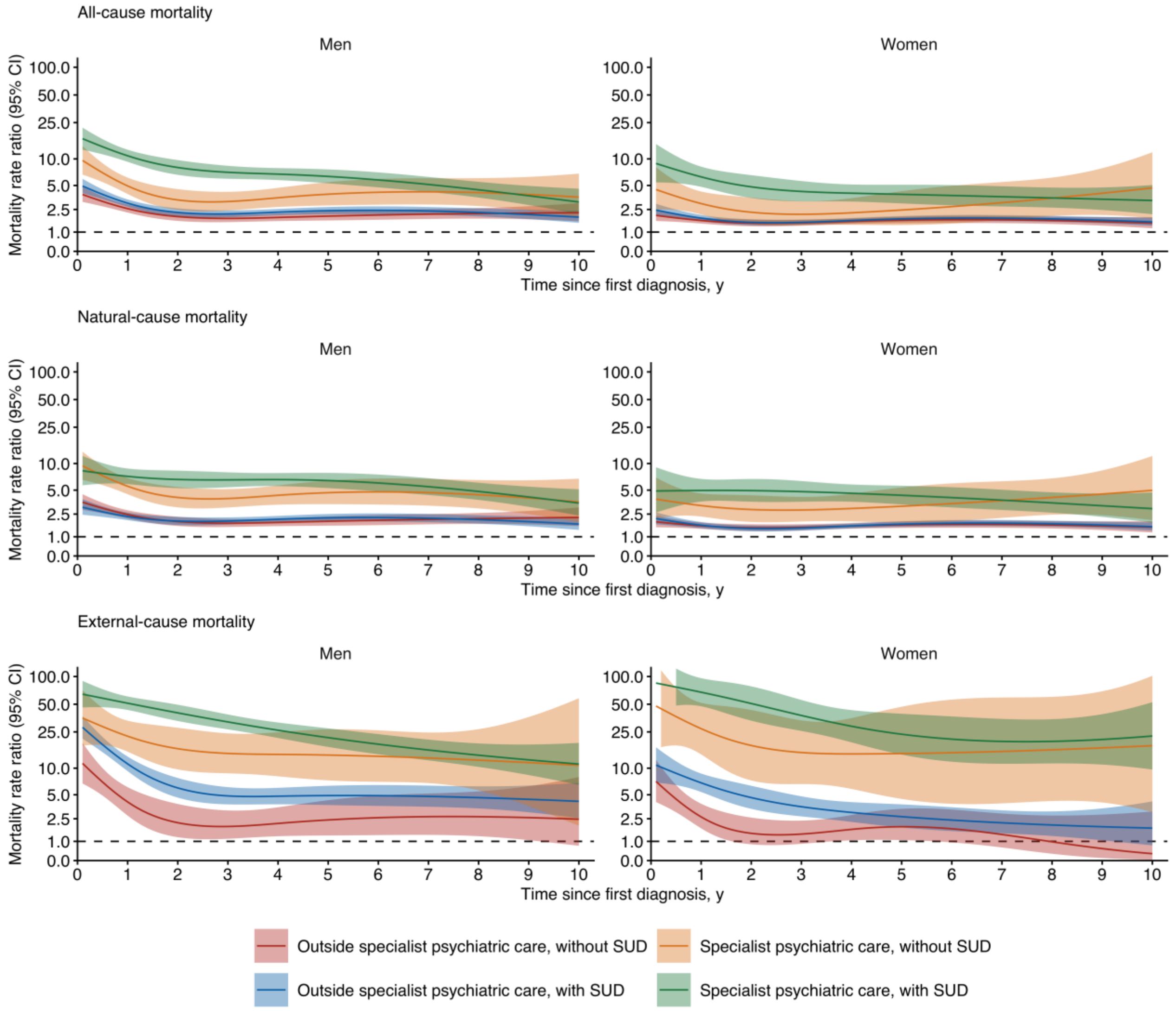

Abbreviations: MRR, mortality rate ratio; SUD, substance use disorder. MRRs are relative to individuals without the specific diagnostic group. Person-time was dynamically allocated according to time-varying psychiatric care setting and SUD status. MRRs were estimated separately for women and men using Poisson regression adjusted for age group, calendar period, region, and Charlson Comorbidity Index. Estimates are shown for the first 10 years after diagnosis. Shaded areas indicate 95% CIs; the dashed horizontal line indicates an MRR of 1. The y-axis is logarithmic.

**Figure S3. Time-varying mortality rate ratios for bipolar or related disorders (6A6) by psychiatric care setting, substance use disorder status, and cause of death**

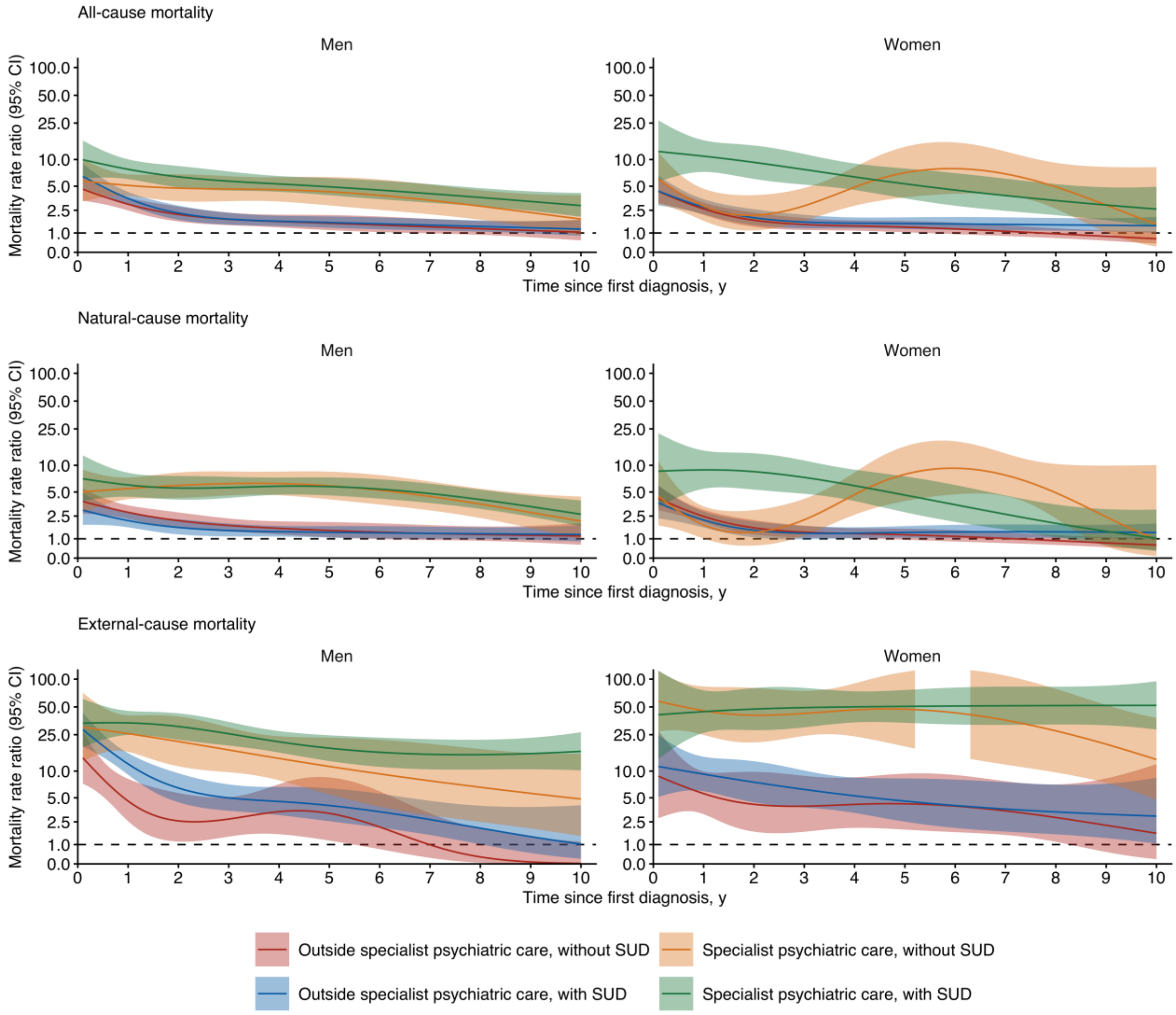

Abbreviations: MRR, mortality rate ratio; SUD, substance use disorder. MRRs are relative to individuals without the specific diagnostic group. Person-time was dynamically allocated according to time-varying psychiatric care setting and SUD status. MRRs were estimated separately for women and men using Poisson regression adjusted for age group, calendar period, region, and Charlson Comorbidity Index. Estimates are shown for the first 10 years after diagnosis. Shaded areas indicate 95% CIs; the dashed horizontal line indicates an MRR of 1. The y-axis is logarithmic.

**Figure S4. Time-varying mortality rate ratios for depressive disorders (6A7) by psychiatric care setting, substance use disorder status, and cause of death**

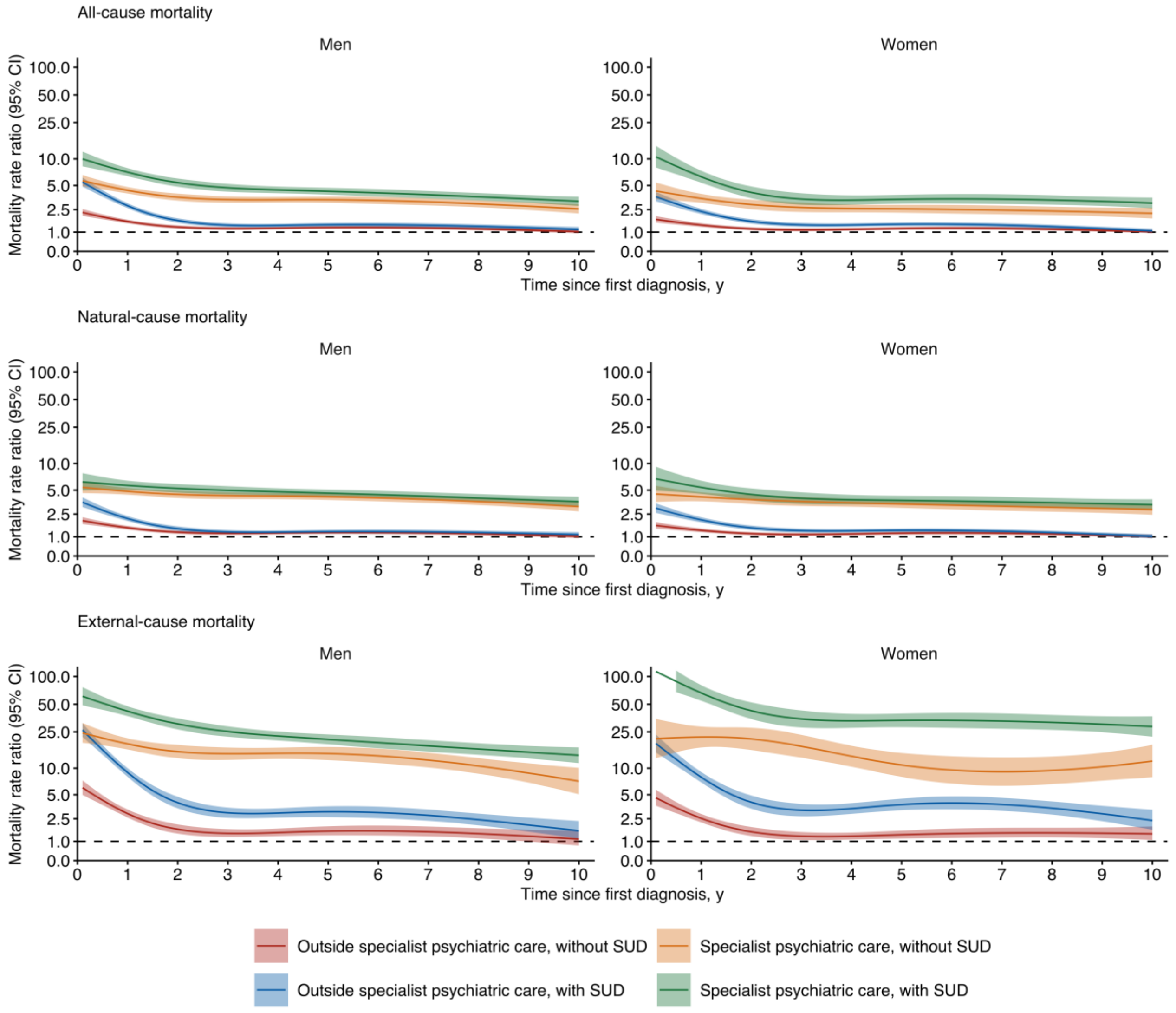

Abbreviations: MRR, mortality rate ratio; SUD, substance use disorder. MRRs are relative to individuals without the specific diagnostic group. Person-time was dynamically allocated according to time-varying psychiatric care setting and SUD status. MRRs were estimated separately for women and men using Poisson regression adjusted for age group, calendar period, region, and Charlson Comorbidity Index. Estimates are shown for the first 10 years after diagnosis. Shaded areas indicate 95% CIs; the dashed horizontal line indicates an MRR of 1. The y-axis is logarithmic.

**Figure S5. Time-varying mortality rate ratios for anxiety or fear-related disorders (6B0) by psychiatric care setting, substance use disorder status, and cause of death**

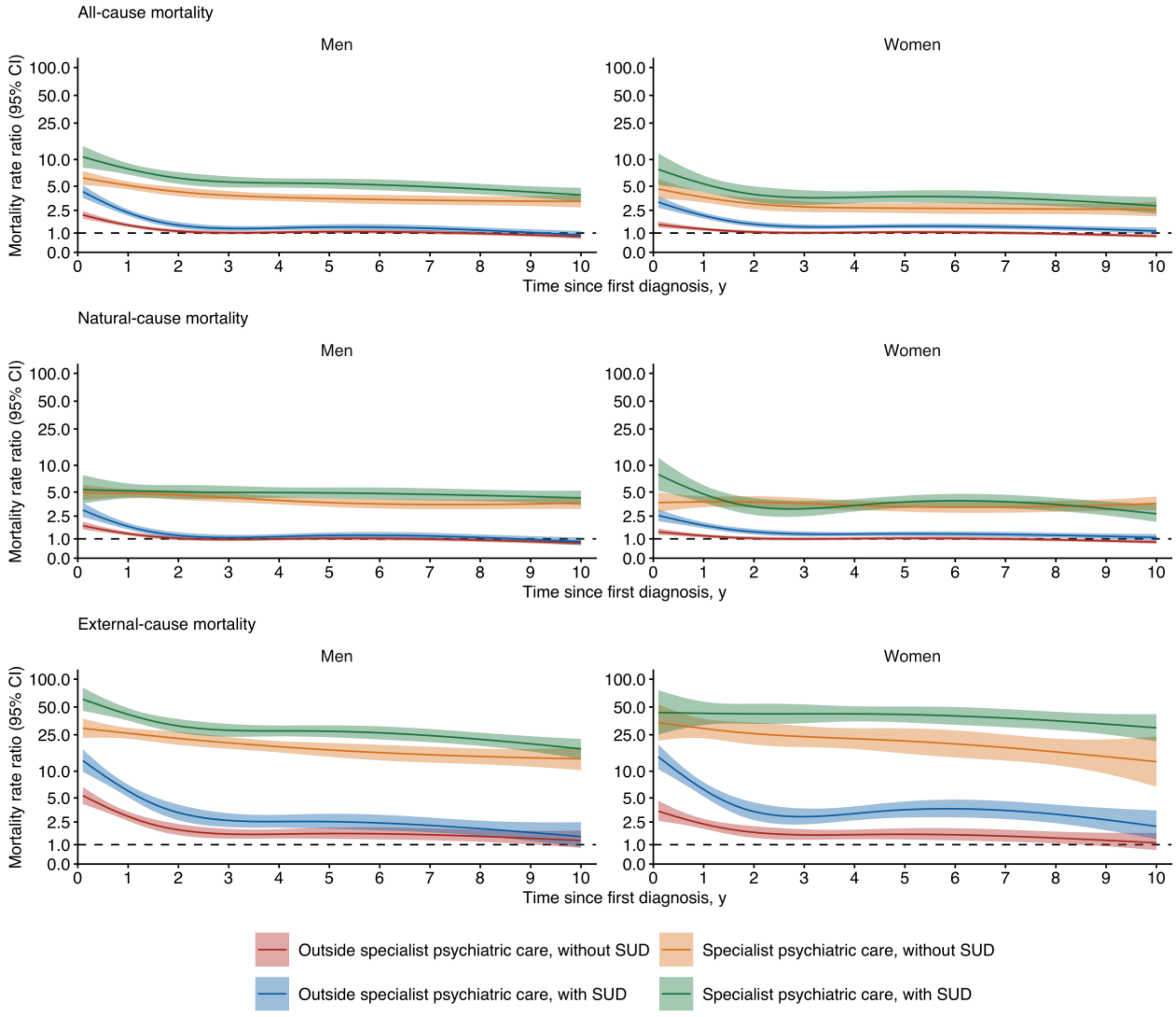

Abbreviations: MRR, mortality rate ratio; SUD, substance use disorder. MRRs are relative to individuals without the specific diagnostic group. Person-time was dynamically allocated according to time-varying psychiatric care setting and SUD status. MRRs were estimated separately for women and men using Poisson regression adjusted for age group, calendar period, region, and Charlson Comorbidity Index. Estimates are shown for the first 10 years after diagnosis. Shaded areas indicate 95% CIs; the dashed horizontal line indicates an MRR of 1. The y-axis is logarithmic.

**Figure S6. Time-varying mortality rate ratios for obsessive-compulsive or related disorders (6B2) by psychiatric care setting, substance use disorder status, and cause of death**

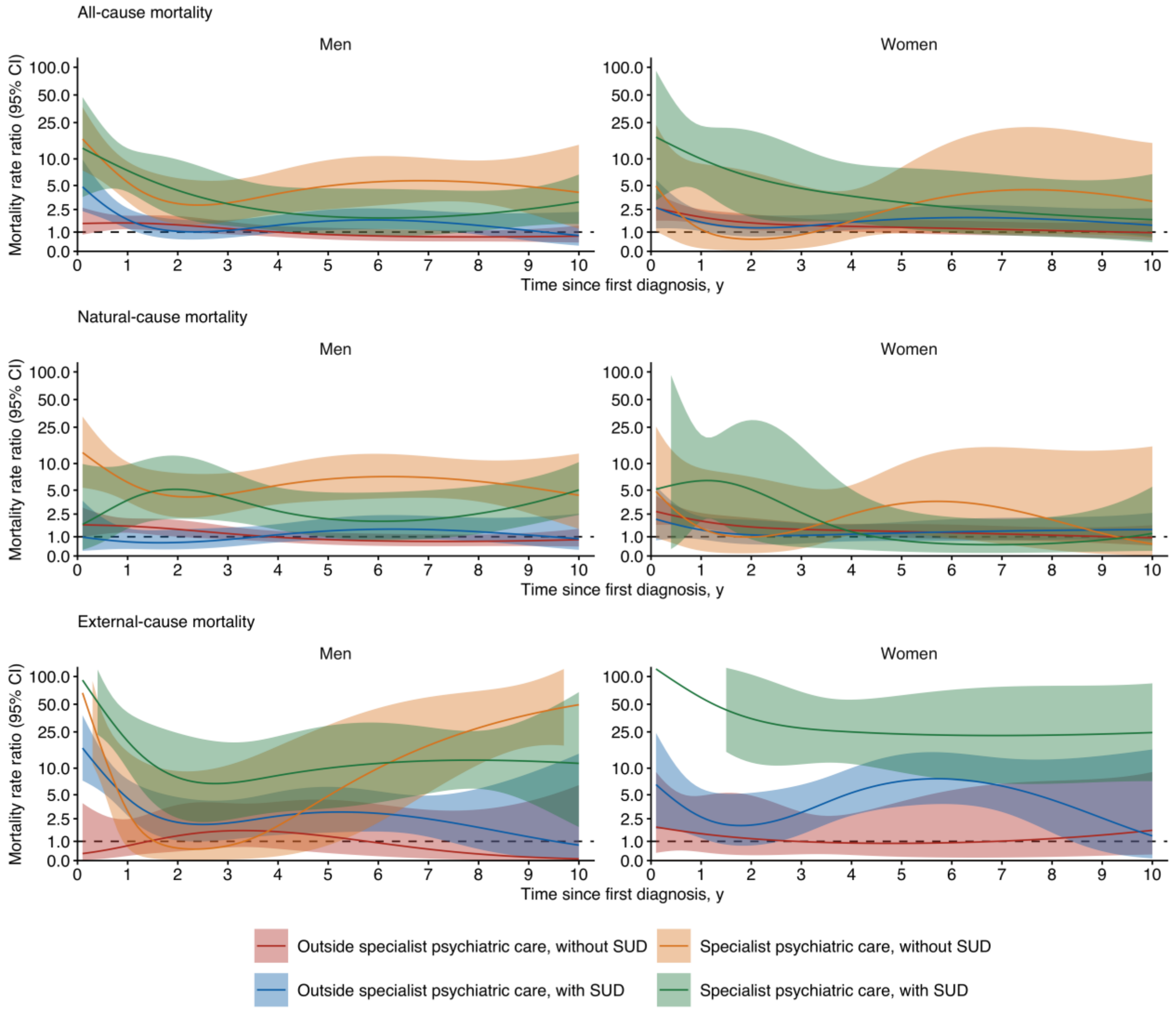

Abbreviations: MRR, mortality rate ratio; SUD, substance use disorder. MRRs are relative to individuals without the specific diagnostic group. Person-time was dynamically allocated according to time-varying psychiatric care setting and SUD status. MRRs were estimated separately for women and men using Poisson regression adjusted for age group, calendar period, region, and Charlson Comorbidity Index. Estimates are shown for the first 10 years after diagnosis. Shaded areas indicate 95% CIs; the dashed horizontal line indicates an MRR of 1. The y-axis is logarithmic.

**Figure S7. Time-varying mortality rate ratios for disorders specifically associated with stress (6B4) by psychiatric care setting, substance use disorder status, and cause of death**

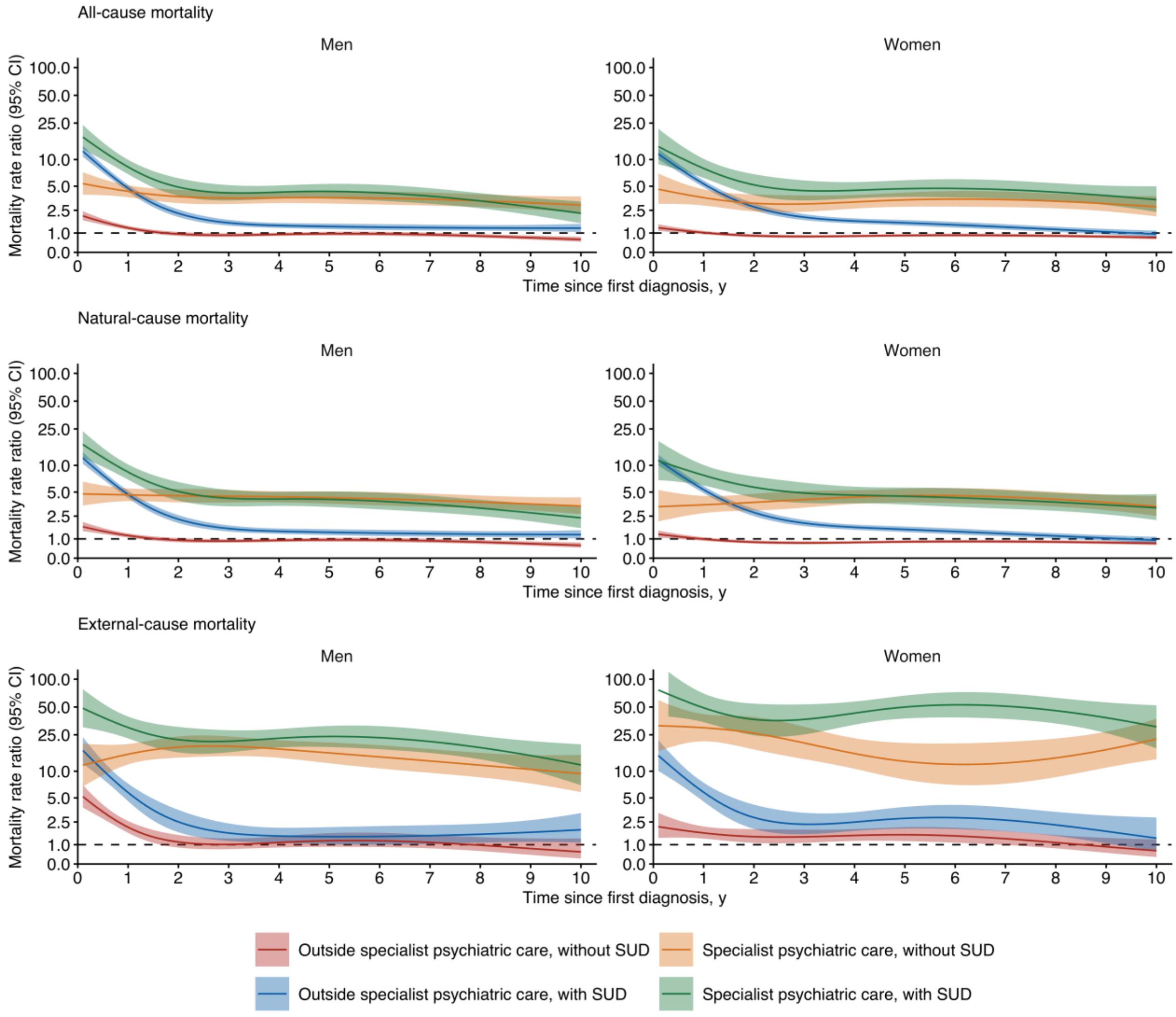

Abbreviations: MRR, mortality rate ratio; SUD, substance use disorder. MRRs are relative to individuals without the specific diagnostic group. Person-time was dynamically allocated according to time-varying psychiatric care setting and SUD status. MRRs were estimated separately for women and men using Poisson regression adjusted for age group, calendar period, region, and Charlson Comorbidity Index. Estimates are shown for the first 10 years after diagnosis. Shaded areas indicate 95% CIs; the dashed horizontal line indicates an MRR of 1. The y-axis is logarithmic.

**Figure S8. Time-varying mortality rate ratios for dissociative disorders (6B6) by psychiatric care setting, substance use disorder status, and cause of death**

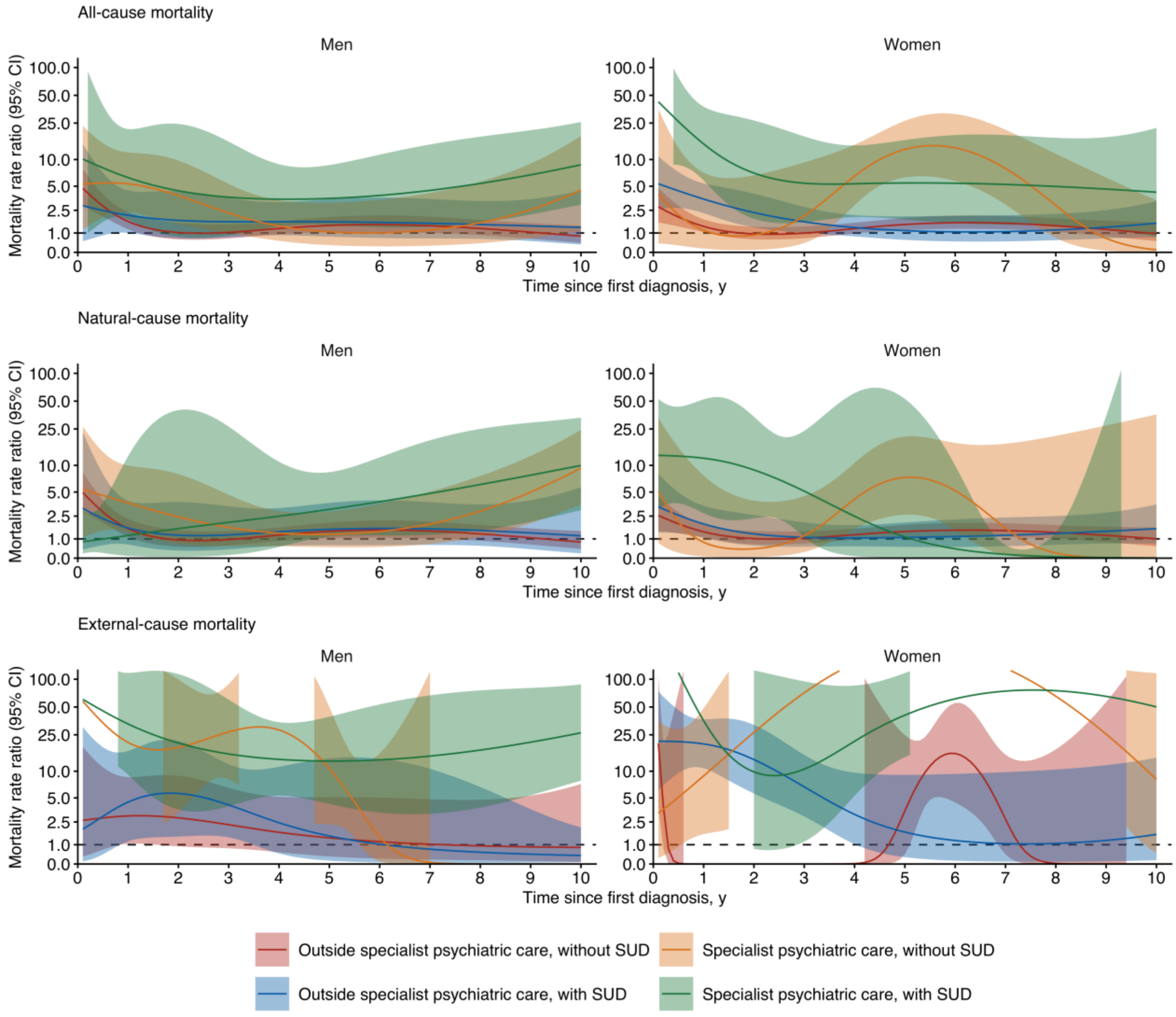

Abbreviations: MRR, mortality rate ratio; SUD, substance use disorder. MRRs are relative to individuals without the specific diagnostic group. Person-time was dynamically allocated according to time-varying psychiatric care setting and SUD status. MRRs were estimated separately for women and men using Poisson regression adjusted for age group, calendar period, region, and Charlson Comorbidity Index. Estimates are shown for the first 10 years after diagnosis. Shaded areas indicate 95% CIs; the dashed horizontal line indicates an MRR of 1. The y-axis is logarithmic.

**Figure S9. Time-varying mortality rate ratios for feeding or eating disorders (6B8) by psychiatric care setting, substance use disorder status, and cause of death**

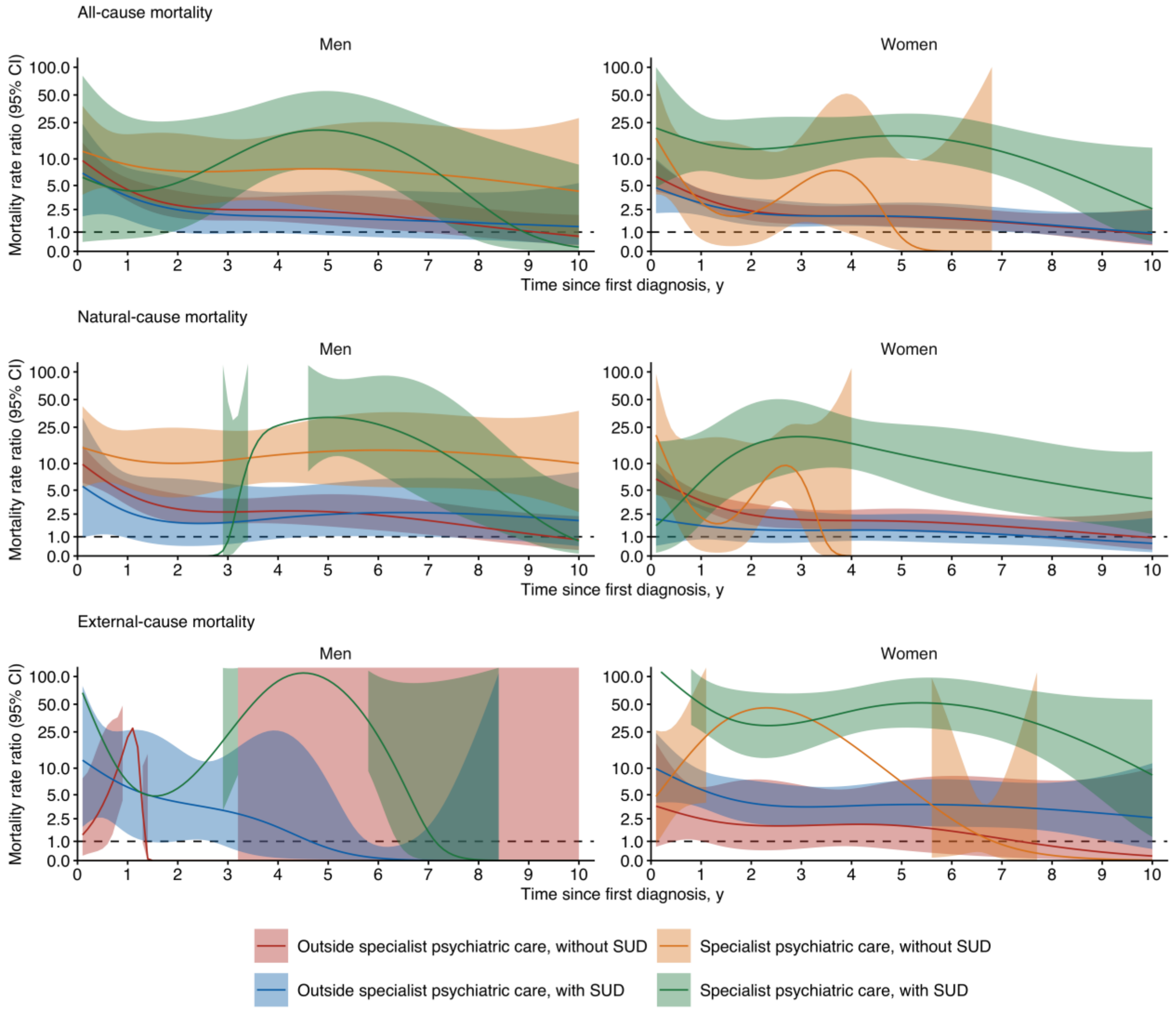

Abbreviations: MRR, mortality rate ratio; SUD, substance use disorder. MRRs are relative to individuals without the specific diagnostic group. Person-time was dynamically allocated according to time-varying psychiatric care setting and SUD status. MRRs were estimated separately for women and men using Poisson regression adjusted for age group, calendar period, region, and Charlson Comorbidity Index. Estimates are shown for the first 10 years after diagnosis. Shaded areas indicate 95% CIs; the dashed horizontal line indicates an MRR of 1. The y-axis is logarithmic.

**Figure S10. Time-varying mortality rate ratios for disorders of bodily distress or bodily experience (6C2) by psychiatric care setting, substance use disorder status, and cause of death**

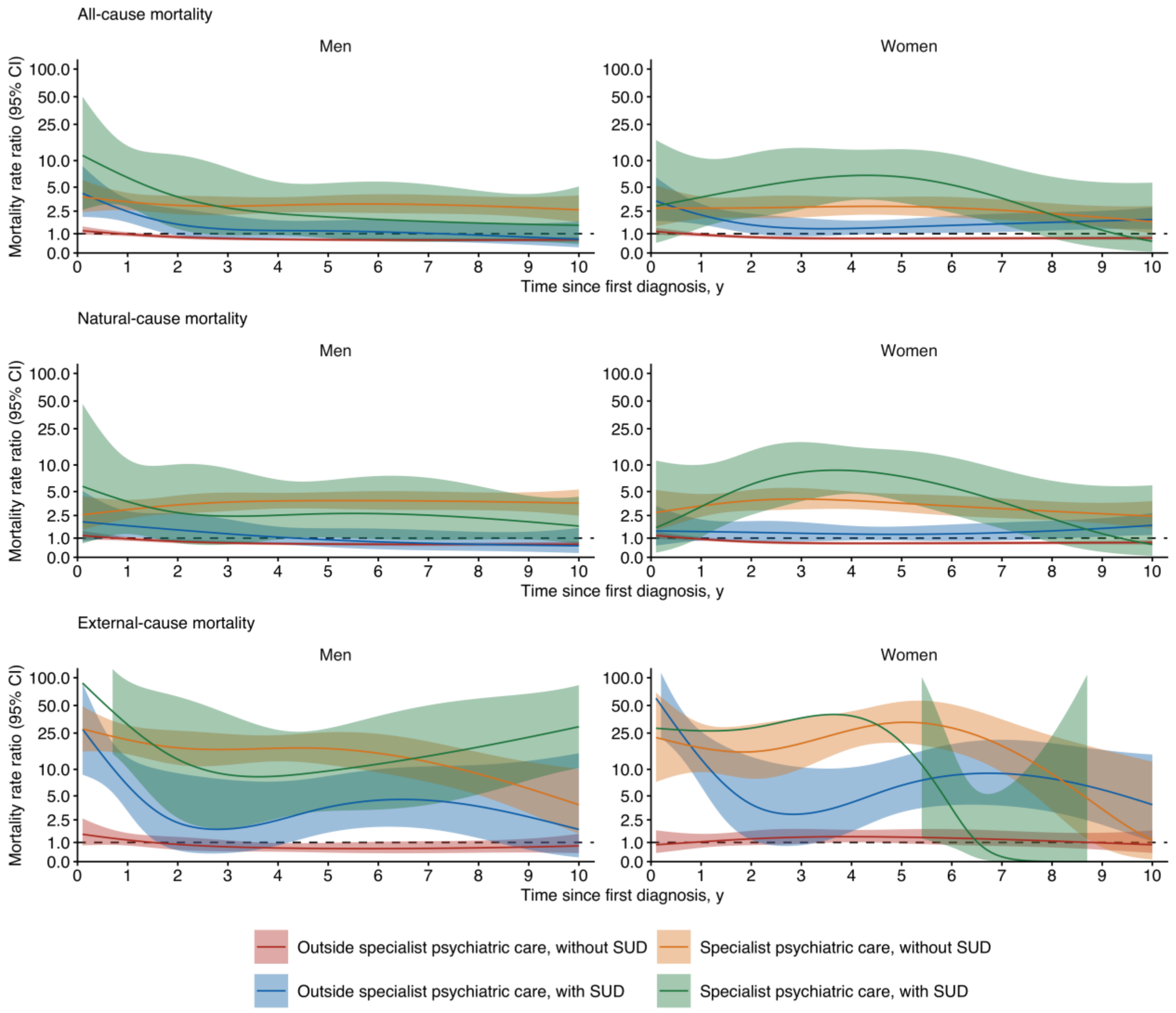

Abbreviations: MRR, mortality rate ratio; SUD, substance use disorder. MRRs are relative to individuals without the specific diagnostic group. Person-time was dynamically allocated according to time-varying psychiatric care setting and SUD status. MRRs were estimated separately for women and men using Poisson regression adjusted for age group, calendar period, region, and Charlson Comorbidity Index. Estimates are shown for the first 10 years after diagnosis. Shaded areas indicate 95% CIs; the dashed horizontal line indicates an MRR of 1. The y-axis is logarithmic.

**Figure S11. Time-varying mortality rate ratios for personality disorders and related traits (6D10-6D11) by psychiatric care setting, substance use disorder status, and cause of death**

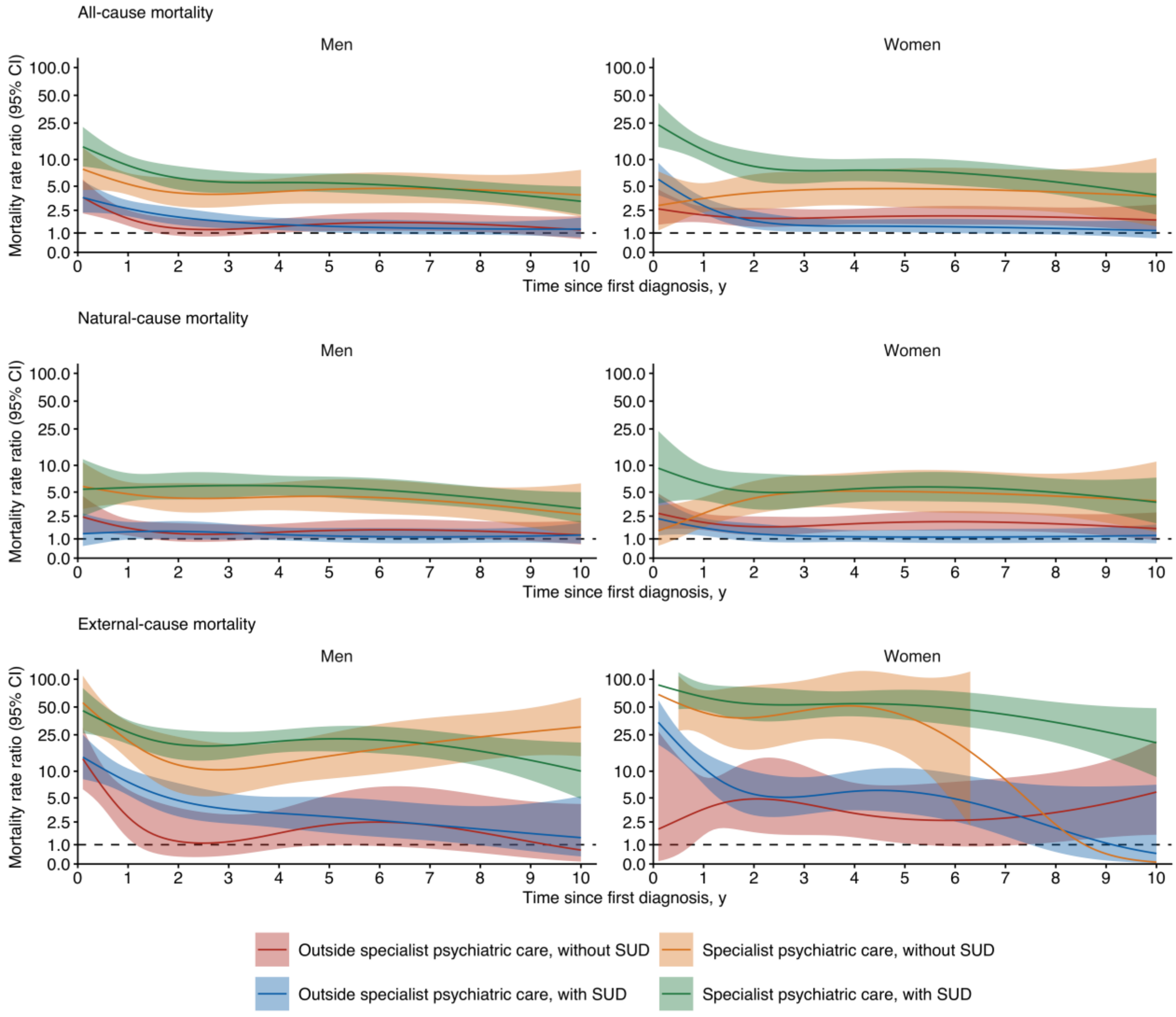

Abbreviations: MRR, mortality rate ratio; SUD, substance use disorder. MRRs are relative to individuals without the specific diagnostic group. Person-time was dynamically allocated according to time-varying psychiatric care setting and SUD status. MRRs were estimated separately for women and men using Poisson regression adjusted for age group, calendar period, region, and Charlson Comorbidity Index. Estimates are shown for the first 10 years after diagnosis. Shaded areas indicate 95% CIs; the dashed horizontal line indicates an MRR of 1. The y-axis is logarithmic.

**Figure S12. Time-varying mortality rate ratios for neurocognitive disorders (6D7-6E0) by psychiatric care setting, substance use disorder status, and cause of death**

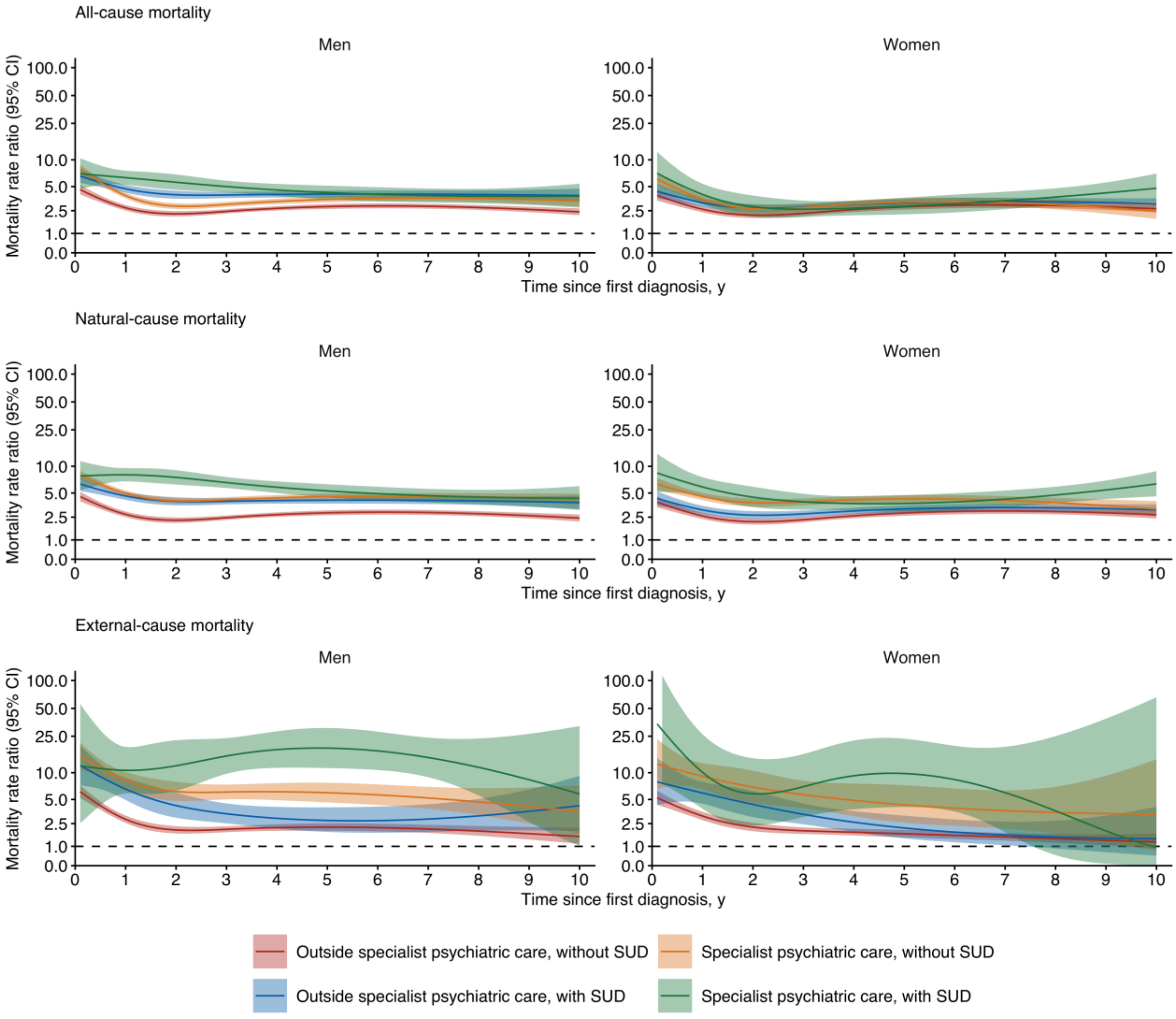

Abbreviations: MRR, mortality rate ratio; SUD, substance use disorder. MRRs are relative to individuals without the specific diagnostic group. Person-time was dynamically allocated according to time-varying psychiatric care setting and SUD status. MRRs were estimated separately for women and men using Poisson regression adjusted for age group, calendar period, region, and Charlson Comorbidity Index. Estimates are shown for the first 10 years after diagnosis. Shaded areas indicate 95% CIs; the dashed horizontal line indicates an MRR of 1. The y-axis is logarithmic.

**Figure S13. Time-varying mortality rate ratios for dementia (6D8) by psychiatric care setting, substance use disorder status, and cause of death**

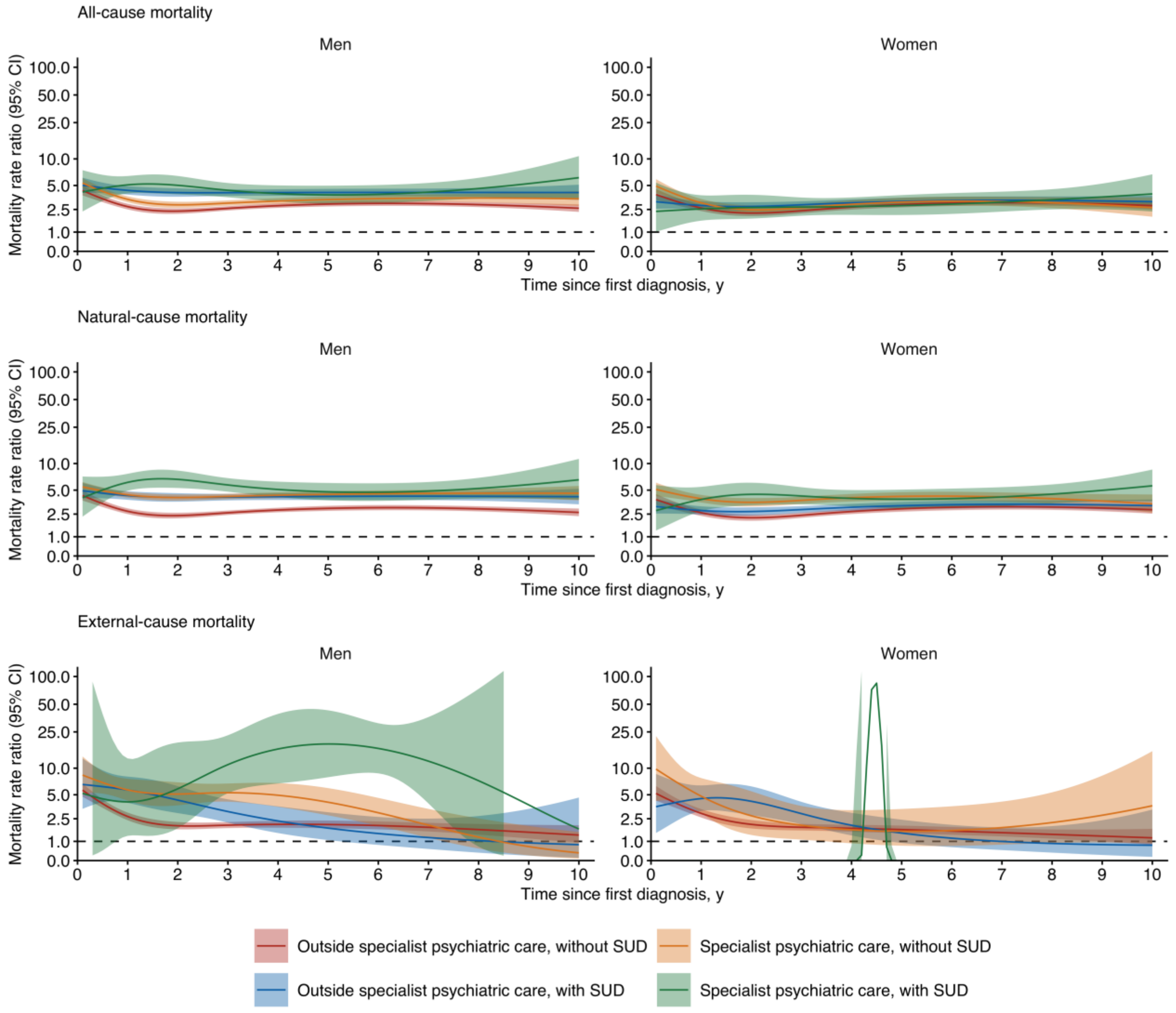

Abbreviations: MRR, mortality rate ratio; SUD, substance use disorder. MRRs are relative to individuals without the specific diagnostic group. Person-time was dynamically allocated according to time-varying psychiatric care setting and SUD status. MRRs were estimated separately for women and men using Poisson regression adjusted for age group, calendar period, region, and Charlson Comorbidity Index. Estimates are shown for the first 10 years after diagnosis. Shaded areas indicate 95% CIs; the dashed horizontal line indicates an MRR of 1. The y-axis is logarithmic.

**Figure S14. Time-varying mortality rate ratios for secondary mental or behavioural syndromes associated with disorders or diseases classified elsewhere (6E6) by psychiatric care setting, substance use disorder status, and cause of death**

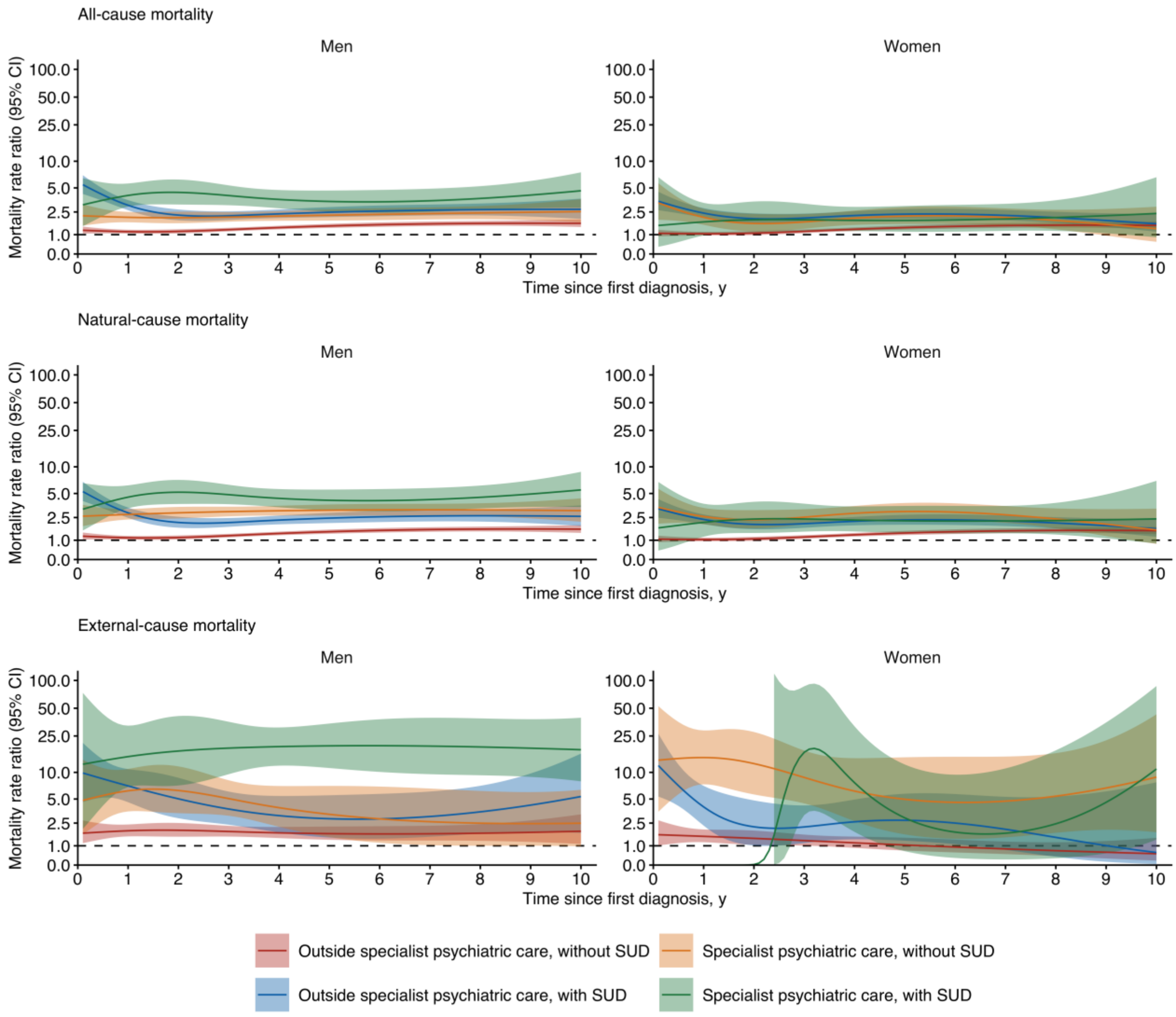

Abbreviations: MRR, mortality rate ratio; SUD, substance use disorder. MRRs are relative to individuals without the specific diagnostic group. Person-time was dynamically allocated according to time-varying psychiatric care setting and SUD status. MRRs were estimated separately for women and men using Poisson regression adjusted for age group, calendar period, region, and Charlson Comorbidity Index. Estimates are shown for the first 10 years after diagnosis. Shaded areas indicate 95% CIs; the dashed horizontal line indicates an MRR of 1. The y-axis is logarithmic.

**Table S8. Ten-year differences in restricted mean survival time by ICD-11 diagnostic groups, gender, and cause of death**

| | **Ten-year difference in restricted mean survival time, years (95% CI)** | | | | | |
|---|---|---|---|---|---|---|
| | **Women** | | | **Men** | | |
| | **All-cause mortality** | **Natural-cause mortality** | **External-cause mortality** | **All-cause mortality** | **Natural-cause mortality** | **External-cause mortality** |
| Mental, behavioural or neurodevelopmental disorders (6) | 0.34 (0.33–0.34) | 0.32 (0.32–0.32) | 0.03 (0.03–0.03) | 0.53 (0.53–0.54) | 0.48 (0.48–0.49) | 0.09 (0.09–0.09) |
| Neurodevelopmental disorders (6A0) | 0.06 (0.06–0.07) | 0.06 (0.05–0.06) | 0.01 (0.01–0.01) | 0.07 (0.06–0.07) | 0.05 (0.05–0.06) | 0.02 (0.02–0.02) |
| Schizophrenia or other primary psychotic disorders (6A2) | 0.34 (0.32–0.37) | 0.31 (0.28–0.33) | 0.08 (0.07–0.09) | 0.44 (0.42–0.46) | 0.32 (0.30–0.33) | 0.26 (0.24–0.29) |
| Bipolar or related disorders (6A6) | 0.09 (0.08–0.11) | 0.06 (0.05–0.07) | 0.06 (0.05–0.07) | 0.28 (0.25–0.30) | 0.16 (0.14–0.18) | 0.17 (0.15–0.20) |
| Depressive disorders (6A7) | 0.08 (0.07–0.08) | 0.06 (0.06–0.06) | 0.02 (0.02–0.02) | 0.18 (0.17–0.18) | 0.12 (0.11–0.13) | 0.09 (0.08–0.09) |
| Anxiety or fear-related disorders (6B0) | 0.03 (0.03–0.04) | 0.02 (0.02–0.03) | 0.02 (0.02–0.02) | 0.09 (0.08–0.09) | 0.04 (0.04–0.05) | 0.06 (0.06–0.06) |
| Obsessive-compulsive or related disorders (6B2) | 0.03 (0.02–0.04) | 0.02 (0.01–0.03) | 0.02 (0.01–0.02) | 0.06 (0.05–0.08) | 0.03 (0.02–0.05) | 0.04 (0.03–0.06) |
| Disorders specifically associated with stress (6B4) | 0.05 (0.05–0.06) | 0.04 (0.04–0.05) | 0.01 (0.01–0.01) | 0.12 (0.11–0.13) | 0.09 (0.08–0.10) | 0.05 (0.04–0.05) |
| Dissociative disorders (6B6) | 0.07 (0.05–0.10) | 0.05 (0.03–0.07) | 0.05 (0.03–0.07) | 0.17 (0.12–0.23) | 0.13 (0.08–0.18) | 0.07 (0.04–0.13) |
| Feeding or eating disorders (6B8) | 0.05 (0.04–0.05) | 0.03 (0.02–0.04) | 0.02 (0.02–0.03) | 0.20 (0.17–0.24) | 0.17 (0.14–0.20) | 0.03 (0.01–0.23) |
| Disorders due to substance use (6C4) | 0.40 (0.39–0.41) | 0.35 (0.34–0.36) | 0.09 (0.08–0.10) | 0.74 (0.73–0.75) | 0.63 (0.61–0.64) | 0.20 (0.20–0.21) |
| Personality disorders and related traits (6D10-6D11) | 0.08 (0.07–0.09) | 0.04 (0.03–0.05) | 0.06 (0.05–0.07) | 0.19 (0.17–0.21) | 0.10 (0.08–0.11) | 0.13 (0.11–0.15) |
| Neurocognitive disorders (6D7-6E0) | 1.57 (1.56–1.59) | 1.55 (1.53–1.56) | 0.08 (0.07–0.09) | 2.04 (2.02–2.06) | 1.99 (1.97–2.02) | 0.18 (0.17–0.19) |

| | Ten-year difference in restricted mean survival time, years (95% CI) | | | | | |
|---|---|---|---|---|---|---|
| | **Women** | | | **Men** | | |
| | **All-cause mortality** | **Natural-cause mortality** | **External-cause mortality** | **All-cause mortality** | **Natural-cause mortality** | **External-cause mortality** |
| Dementia (6D8) | 1.65 (1.64–1.67) | 1.63 (1.61–1.65) | 0.08 (0.07–0.08) | 2.15 (2.13–2.18) | 2.11 (2.09–2.14) | 0.17 (0.15–0.18) |

Abbreviations: RMST, restricted mean survival time.

RMST differences represent the difference in model-predicted survival during the first 10 years after diagnosis between individuals with and without the specific diagnostic group. Estimates were obtained separately for women and men using Poisson regression allowing mortality rates to vary with time since diagnosis and adjusted for age group, calendar period, region, and Charlson Comorbidity Index. Natural and external causes of death are competing events; therefore, cause-specific RMST differences do not sum to the overall RMST difference.

**Table S9. Ten-year differences in restricted mean survival time by psychiatric care setting and substance use disorder status across diagnostic groups**

| | **Ten-year difference in restricted mean survival time, years (95% CI)** | | | | | | | |
|---|---|---|---|---|---|---|---|---|
| | **Men** | | | | **Women** | | | |
| | **Outside specialist psychiatric care** | | **Specialist psychiatric care** | | **Outside specialist psychiatric care** | | **Specialist psychiatric care** | |
| | **No SUD** | **SUD** | **No SUD** | **SUD** | **No SUD** | **SUD** | **No SUD** | **SUD** |
| Mental, behavioural or neurodevelopmental disorders (6) | 0.45 (0.45–0.46) | 0.74 (0.73–0.75) | 0.62 (0.60–0.63) | 1.17 (1.13–1.20) | 0.32 (0.31–0.32) | 0.54 (0.52–0.56) | 0.41 (0.40–0.42) | 0.79 (0.74–0.84) |
| Neurodevelopmental disorders (6A0) | 0.06 (0.06–0.07) | 0.11 (0.09–0.13) | 0.04 (0.03–0.05) | 0.25 (0.22–0.29) | 0.06 (0.06–0.07) | 0.11 (0.08–0.14) | 0.05 (0.04–0.07) | 0.25 (0.19–0.34) |
| Schizophrenia or other primary psychotic disorders (6A2) | 0.29 (0.25–0.32) | 0.68 (0.58–0.77) | 0.39 (0.37–0.42) | 1.14 (1.07–1.21) | 0.29 (0.26–0.33) | 0.70 (0.51–0.91) | 0.34 (0.31–0.37) | 1.17 (1.02–1.32) |
| Bipolar or related disorders (6A6) | 0.17 (0.13–0.21) | 0.48 (0.38–0.61) | 0.22 (0.18–0.26) | 0.63 (0.54–0.72) | 0.07 (0.05–0.09) | 0.24 (0.17–0.33) | 0.08 (0.06–0.10) | 0.32 (0.26–0.40) |
| Depressive disorders (6A7) | 0.09 (0.08–0.09) | 0.46 (0.43–0.48) | 0.20 (0.19–0.21) | 0.67 (0.63–0.70) | 0.05 (0.04–0.05) | 0.26 (0.24–0.29) | 0.12 (0.11–0.13) | 0.43 (0.39–0.46) |
| Anxiety or fear-related disorders (6B0) | 0.03 (0.02–0.03) | 0.30 (0.28–0.32) | 0.08 (0.07–0.09) | 0.45 (0.42–0.49) | 0.01 (0.01–0.02) | 0.18 (0.16–0.20) | 0.07 (0.06–0.07) | 0.27 (0.24–0.30) |
| Obsessive-compulsive or related disorders (6B2) | 0.02 (0.00–0.04) | 0.32 (0.23–0.44) | 0.07 (0.05–0.11) | 0.28 (0.18–0.44) | 0.03 (0.02–0.04) | 0.06 (0.01–0.20) | 0.04 (0.02–0.06) | 0.19 (0.11–0.33) |
| Disorders specifically associated with stress (6B4) | 0.01 (0.00–0.02) | 0.41 (0.37–0.46) | 0.25 (0.23–0.27) | 0.59 (0.52–0.66) | -0.01 (-0.01–-0.00) | 0.25 (0.22–0.29) | 0.19 (0.18–0.20) | 0.39 (0.33–0.45) |
| Dissociative disorders (6B6) | 0.14 (0.08–0.21) | 0.37 (0.16–0.72) | 0.17 (0.06–0.36) | 0.68 (0.36–1.44) | 0.05 (0.02–0.07) | 0.32 (0.20–0.67) | 0.11 (0.07–0.18) | 0.63 (0.39–1.07) |

| | Ten-year difference in restricted mean survival time, years (95% CI) | | | | | | | |
|---|---|---|---|---|---|---|---|---|
| | **Men** | | | | **Women** | | | |
| | **Outside specialist psychiatric care** | | **Specialist psychiatric care** | | **Outside specialist psychiatric care** | | **Specialist psychiatric care** | |
| | **No SUD** | **SUD** | **No SUD** | **SUD** | **No SUD** | **SUD** | **No SUD** | **SUD** |
| Feeding or eating disorders (6B8) | 0.18 (0.14–0.23) | 0.41 (0.27–0.66) | 0.15 (0.09–0.27) | 0.50 (0.30–1.21) | 0.04 (0.03–0.05) | 0.06 (0.03–2.77) | 0.05 (0.03–0.06) | 0.27 (0.20–0.39) |
| Personality disorders and related traits (6D10-6D11) | 0.08 (0.04–0.12) | 0.38 (0.30–0.48) | 0.10 (0.08–0.14) | 0.54 (0.46–0.62) | 0.05 (0.03–0.06) | 0.14 (0.10–0.21) | 0.05 (0.04–0.07) | 0.35 (0.29–0.41) |
| Neurocognitive disorders (6D7-6E0) | 1.95 (1.93–1.97) | 2.66 (2.58–2.73) | 2.93 (2.85–3.00) | 3.30 (3.06–3.55) | 1.55 (1.53–1.57) | 2.02 (1.91–2.13) | 1.86 (1.80–1.92) | 2.14 (1.79–2.51) |
| Dementia (6D8) | 2.09 (2.07–2.11) | 2.51 (2.42–2.60) | 2.95 (2.86–3.04) | 3.02 (2.72–3.32) | 1.64 (1.62–1.66) | 1.91 (1.79–2.03) | 1.80 (1.73–1.87) | 1.63 (1.27–2.10) |

Abbreviation: RMST, restricted mean survival time. RMST differences represent the difference in model-predicted survival during the first 10 years after diagnosis between individuals with and without the specific diagnostic group. Estimates were obtained separately for women and men using Poisson regression allowing mortality rates to vary with time since diagnosis and adjusted for age group, calendar period, region, and Charlson Comorbidity Index. Positive values indicate shorter predicted survival among individuals with the diagnosis; negative values indicate longer predicted survival.

**Table S10. Estimated ten-year restricted mean survival time by diagnostic group, gender, and cause of death**

| | | Estimated 10-year restricted mean survival time, years | | | | | |
|---|---|---|---|---|---|---|---|
| | | With diagnosis | | | Without diagnosis | | |
| | | All-cause mortality | Natural-cause mortality | External-cause mortality | All-cause mortality | Natural-cause mortality | External-cause mortality |
| Mental, behavioural or neurodevelopmental disorders (6) | Men | 8.93 | 8.86 | 9.74 | 9.41 | 9.39 | 9.83 |
| | Women | 9.07 | 9.04 | 9.69 | 9.39 | 9.38 | 9.72 |
| Neurodevelopmental disorders (6A0) | Men | 9.89 | 9.87 | 9.97 | 9.95 | 9.94 | 9.98 |
| | Women | 9.86 | 9.85 | 9.96 | 9.92 | 9.92 | 9.97 |
| Schizophrenia or other primary psychotic disorders (6A2) | Men | 9.23 | 9.08 | 9.63 | 9.55 | 9.52 | 9.90 |
| | Women | 8.73 | 8.67 | 9.58 | 9.04 | 9.02 | 9.66 |
| Bipolar or related disorders (6A6) | Men | 9.64 | 9.51 | 9.79 | 9.80 | 9.78 | 9.96 |
| | Women | 9.80 | 9.76 | 9.90 | 9.86 | 9.85 | 9.96 |
| Depressive disorders (6A7) | Men | 9.59 | 9.51 | 9.85 | 9.71 | 9.69 | 9.94 |
| | Women | 9.67 | 9.65 | 9.90 | 9.73 | 9.72 | 9.93 |
| Anxiety or fear-related disorders (6B0) | Men | 9.82 | 9.77 | 9.91 | 9.87 | 9.85 | 9.97 |
| | Women | 9.83 | 9.82 | 9.95 | 9.86 | 9.85 | 9.97 |
| Obsessive-compulsive or related disorders (6B2) | Men | 9.86 | 9.82 | 9.93 | 9.90 | 9.88 | 9.97 |
| | Women | 9.92 | 9.90 | 9.97 | 9.94 | 9.93 | 9.98 |

| | | Estimated 10-year restricted mean survival time, years | | | | | |
|---|---|---|---|---|---|---|---|
| | | **With diagnosis** | | | **Without diagnosis** | | |
| | | **All-cause mortality** | **Natural-cause mortality** | **External-cause mortality** | **All-cause mortality** | **Natural-cause mortality** | **External-cause mortality** |
| Disorders specifically associated with stress (6B4) | Men | 9.72 | 9.66 | 9.92 | 9.81 | 9.78 | 9.97 |
| | Women | 9.82 | 9.80 | 9.97 | 9.86 | 9.86 | 9.98 |
| Dissociative disorders (6B6) | Men | 9.56 | 9.49 | 9.86 | 9.69 | 9.66 | 9.94 |
| | Women | 9.75 | 9.72 | 9.89 | 9.80 | 9.79 | 9.94 |
| Feeding or eating disorders (6B8) | Men | 9.68 | 9.64 | 9.93 | 9.86 | 9.84 | 9.96 |
| | Women | 9.93 | 9.91 | 9.96 | 9.96 | 9.96 | 9.98 |
| Disorders due to substance use (6C4) | Men | 8.92 | 8.78 | 9.75 | 9.55 | 9.53 | 9.95 |
| | Women | 9.31 | 9.25 | 9.85 | 9.66 | 9.65 | 9.94 |
| Personality disorders and related traits (6D10-6D11) | Men | 9.78 | 9.67 | 9.84 | 9.88 | 9.86 | 9.97 |
| | Women | 9.90 | 9.86 | 9.93 | 9.94 | 9.94 | 9.99 |
| Neurocognitive disorders (6D7-6E0) | Men | 5.07 | 4.95 | 8.80 | 7.06 | 6.99 | 8.98 |
| | Women | 5.39 | 5.32 | 8.33 | 6.93 | 6.89 | 8.41 |
| Dementia (6D8) | Men | 4.67 | 4.55 | 8.69 | 6.78 | 6.70 | 8.86 |
| | Women | 5.11 | 5.05 | 8.22 | 6.74 | 6.70 | 8.30 |

Abbreviation: RMST, restricted mean survival time. RMST was estimated separately for women and men from model-predicted survival using Poisson regression allowing mortality rates to vary with time since diagnosis and adjusted for age group, calendar period, region, and Charlson Comorbidity Index.

Differences in RMST between individuals with and without the specific diagnostic group represent predicted survival differences during the first 10 years after diagnosis. Overall RMST reflects all-cause mortality. Natural- and external-cause RMSTs were estimated with deaths from other causes treated as competing events and therefore do not sum to overall RMST.

**Table S11. Estimated ten-year restricted mean survival time by diagnostic group, gender, psychiatric care setting, and substance use disorder status**

| | | Estimated 10-year restricted mean survival time, years | | | | |
|---|---|---|---|---|---|---|
| | | With diagnosis | | | | Without diagnosis |
| | | Outside specialist psychiatric care | | Specialist psychiatric care | | |
| | | No SUD | SUD | No SUD | SUD | |
| Mental, behavioural or neurodevelopmental disorders (6) | Men | 8.94 | 8.64 | 8.77 | 8.22 | 9.39 |
| | Women | 9.06 | 8.84 | 8.97 | 8.59 | 9.38 |
| Neurodevelopmental disorders (6A0) | Men | 9.88 | 9.83 | 9.90 | 9.69 | 9.94 |
| | Women | 9.85 | 9.81 | 9.86 | 9.66 | 9.92 |
| Schizophrenia or other primary psychotic disorders (6A2) | Men | 9.24 | 8.85 | 9.13 | 8.38 | 9.52 |
| | Women | 8.72 | 8.32 | 8.68 | 7.85 | 9.02 |
| Bipolar or related disorders (6A6) | Men | 9.62 | 9.30 | 9.56 | 9.16 | 9.78 |
| | Women | 9.78 | 9.61 | 9.78 | 9.53 | 9.85 |
| Depressive disorders (6A7) | Men | 9.60 | 9.23 | 9.49 | 9.02 | 9.69 |
| | Women | 9.68 | 9.46 | 9.60 | 9.30 | 9.72 |
| Anxiety or fear-related disorders (6B0) | Men | 9.83 | 9.56 | 9.77 | 9.40 | 9.85 |
| | Women | 9.84 | 9.67 | 9.78 | 9.58 | 9.85 |
| Obsessive-compulsive or related disorders (6B2) | Men | 9.86 | 9.56 | 9.81 | 9.60 | 9.88 |
| | Women | 9.91 | 9.88 | 9.90 | 9.75 | 9.93 |

| | | Estimated 10-year restricted mean survival time, years | | | | |
|---|---|---|---|---|---|---|
| | | With diagnosis | | | | Without diagnosis |
| | | Outside specialist psychiatric care | | Specialist psychiatric care | | |
| | | No SUD | SUD | No SUD | SUD | |
| Disorders specifically associated with stress (6B4) | Men | 9.77 | 9.37 | 9.54 | 9.20 | 9.78 |
| | Women | 9.86 | 9.60 | 9.67 | 9.47 | 9.86 |
| Dissociative disorders (6B6) | Men | 9.53 | 9.29 | 9.49 | 8.98 | 9.66 |
| | Women | 9.75 | 9.47 | 9.68 | 9.16 | 9.79 |
| Feeding or eating disorders (6B8) | Men | 9.67 | 9.44 | 9.69 | 9.34 | 9.84 |
| | Women | 9.92 | 9.90 | 9.91 | 9.69 | 9.96 |
| Personality disorders and related traits (6D10-6D11) | Men | 9.78 | 9.48 | 9.76 | 9.33 | 9.86 |
| | Women | 9.89 | 9.79 | 9.88 | 9.59 | 9.94 |
| Neurocognitive disorders (6D7-6E0) | Men | 5.03 | 4.33 | 4.06 | 3.68 | 6.99 |
| | Women | 5.34 | 4.87 | 5.03 | 4.75 | 6.89 |
| Dementia (6D8) | Men | 4.61 | 4.19 | 3.75 | 3.68 | 6.70 |
| | Women | 5.06 | 4.79 | 4.89 | 5.07 | 6.70 |

RMST was estimated separately for women and men from model-predicted survival using Poisson regression allowing mortality rates to vary with time since diagnosis and adjusted for age group, calendar period, region, and Charlson Comorbidity Index. Differences in RMST between individuals with and without the specific diagnostic group represent predicted survival differences during the first 10 years after diagnosis. Psychiatric care setting and substance use disorder (SUD) status were time-varying characteristics. The same no-diagnosis reference was used across psychiatric care setting and SUD strata.

**Figure S15. Mortality rate ratios over time since first diagnosis of mental disorders using a stricter incident-cohort definition**

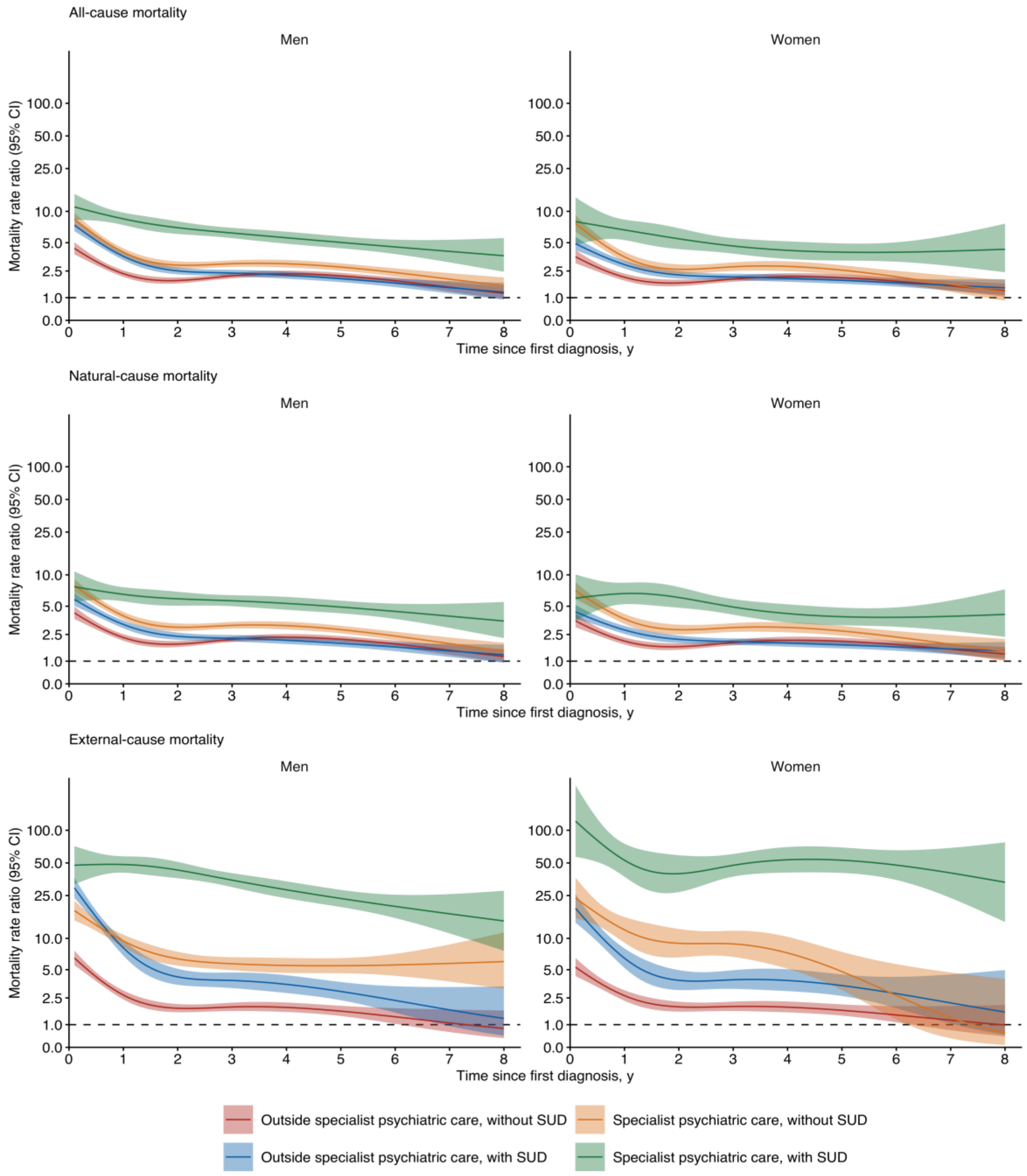

Abbreviations: MRR, mortality rate ratio; SUD, substance use disorder. MRRs are relative to individuals without a diagnosed mental, behavioural, or neurodevelopmental disorder. To reduce potential misclassification of prevalent disorders during the early years of primary care data availability, follow-up was restricted to January 1, 2016, onward. Person-time was dynamically allocated according to time-varying psychiatric care setting and SUD status. MRRs were estimated separately for women and men using Poisson regression adjusted for age group, calendar period, region, and Charlson Comorbidity Index. Shaded areas indicate 95% CIs; the dashed horizontal line indicates an MRR of 1. The y-axis is logarithmic.

**Table S12. Mortality rates and mortality rate ratios associated with ICD-10 mental and behavioural disorder groups**

| | No. (%) | Person-years | No. of deaths | Mortality rate per 1000 person-years | | | Mortality rate ratio (95% CI) | |
|---|---|---|---|---|---|---|---|---|
| **dg_name** | | | | **Crude** | **Age-standardized** | **Population without the diagnosis** | **Basic model** | **CCI-adjusted model** |
| Mental or behavioural disorders (F00-F99) | 1 762 878 (32.0) | 9 325 074 | 180 652 | 19.4 | 15.8 (15.7-15.9) | 6.0 | 2.57 (2.41-2.73) | 2.29 (2.19-2.40) |
| Organic mental disorders (F00–F09) | 218 367 (4.0) | 766 864 | 119 229 | 155.5 | 37.5 (36.6-38.9) | 6.2 | 3.13 (2.93-3.34) | 2.73 (2.58-2.88) |
| Substance use disorders (F10–F19) | 205 890 (3.7) | 1 151 897 | 29 313 | 25.4 | 25.6 (25.3-26.2) | 7.7 | 3.58 (3.29-3.89) | 3.05 (2.87-3.23) |
| Schizophrenia spectrum (F20–F29) | 39 183 (0.7) | 212 212 | 6 313 | 29.7 | 20.9 (20.2-22.1) | 7.9 | 2.34 (2.18-2.52) | 2.17 (2.03-2.31) |
| Mood disorders (F30–F39) | 495 541 (9.0) | 2 646 974 | 27 577 | 10.4 | 12.8 (12.7-13.0) | 7.9 | 1.77 (1.65-1.89) | 1.57 (1.51-1.65) |
| Neurotic, stress–related and somatoform disorders (F40–F48) | 818 616 (14.8) | 4 035 091 | 22 436 | 5.6 | 9.6 (9.5-9.8) | 8.2 | 1.42 (1.31-1.54) | 1.31 (1.24-1.38) |
| Behavioral syndromes (F50–F59) | 528 071 (9.6) | 2 213 501 | 22 329 | 10.1 | 8.3 (8.2-8.5) | 7.9 | 1.02 (0.94-1.11) | 0.98 (0.93-1.02) |
| Personality disorders (F60–F69) | 47 987 (0.9) | 237 723 | 1 257 | 5.3 | 18.1 (16.8-19.5) | 8.0 | 2.65 (2.42-2.90) | 2.38 (2.21-2.56) |
| Intellectual disability (F70–F79) | 17 869 (0.3) | 104 353 | 1 764 | 16.9 | 30.5 (29.0-32.1) | 8.0 | 4.46 (4.08-4.88) | 4.66 (4.34-5.00) |

| | No. (%) | Person-years | No. of deaths | Mortality rate per 1000 person-years | | | Mortality rate ratio (95% CI) | |
|---|---|---|---|---|---|---|---|---|
| dg_name | | | | Crude | Age-standardized | Population without the diagnosis | Basic model | CCI-adjusted model |
| Disorders of psychological development (F80–F89) | 151 637 (2.7) | 843 147 | 1 515 | 1.8 | 22.0 (20.7-23.3) | 8.1 | 2.28 (2.07-2.52) | 1.86 (1.72-2.02) |
| Behavioral and emotional disorders (F90–F98) | 291 835 (5.3) | 1 477 007 | 2 705 | 1.8 | 15.8 (15.1-16.5) | 8.2 | 2.14 (1.98-2.31) | 1.87 (1.75-2.01) |
| Secondary care only: Mental or behavioural disorders | 408 179 (7.4) | 2 416 023 | 23 807 | 9.9 | 18.9 (18.6-19.1) | 6.0 | 3.46 (3.22-3.72) | 2.94 (2.79-3.10) |

MRRs are not shown for diagnostic groups with insufficient deaths for stable estimation. The final row is restricted to diagnoses recorded in specialist psychiatric care. Crude MRs were calculated from deaths and person-years. Age-standardized MRs were estimated by direct standardization to the age distribution of the study population. MRs for the population without the diagnosis are crude rates among individuals without the specific diagnosis. MRRs were estimated using Poisson regression adjusted for gender, age group, calendar period, and region (basic model), with additional adjustment for CCI (CCI-adjusted model). Diagnosis-specific MRRs compare individuals with each diagnosis with individuals without that diagnosis, regardless of other mental disorder diagnoses.

**Table S13. ICD-10 diagnosis-specific mortality rate ratios by psychiatric care setting and substance use disorder status among individuals aged 5 to 64 years**

| | Men | | | | Women | | | |
|---|---|---|---|---|---|---|---|---|
| | **Outside specialist psychiatric care** | | **Specialist psychiatric care** | | **Outside specialist psychiatric care** | | **Specialist psychiatric care** | |
| | **No SUD** | **SUD** | **No SUD** | **SUD** | **No SUD** | **SUD** | **No SUD** | **SUD** |
| Mental or behavioural disorders (F00-F99) | 1.76 (1.60–1.93) | 6.10 (5.41–6.87) | 3.13 (2.82–3.48) | 9.03 (7.92–10.30) | 1.41 (1.27–1.56) | 4.80 (4.00–5.75) | 2.79 (2.49–3.12) | 7.12 (5.95–8.53) |
| Organic mental disorders (F00–F09) | 4.97 (4.31–5.73) | 8.10 (6.63–9.89) | 5.06 (4.09–6.26) | 7.57 (5.94–9.66) | 6.28 (5.18–7.61) | 7.60 (5.42–10.66) | 7.12 (5.51–9.20) | 7.14 (4.46–11.42) |
| Substance use disorders (F10–F19) | NA | 5.47 (4.89–6.13) | 9.16 (2.14–39.25) | 8.83 (7.74–10.07) | NA | 4.17 (3.55–4.91) | NA | 7.17 (5.96–8.64) |
| Schizophrenia spectrum (F20–F29) | 3.36 (2.68–4.21) | 10.58 (8.24–13.58) | 4.45 (3.79–5.23) | 12.48 (10.53–14.79) | 2.52 (1.69–3.76) | 10.76 (6.92–16.72) | 3.74 (3.10–4.52) | 11.27 (8.49–14.96) |
| Mood disorders (F30–F39) | 1.32 (1.19–1.46) | 5.60 (4.95–6.34) | 1.92 (1.69–2.17) | 5.92 (5.10–6.86) | 1.09 (0.98–1.21) | 4.44 (3.67–5.38) | 1.68 (1.50–1.88) | 5.49 (4.51–6.68) |
| Neurotic, stress–related and somatoform disorders (F40–F48) | 1.02 (0.93–1.13) | 5.45 (4.79–6.21) | 2.52 (2.25–2.81) | 7.04 (6.10–8.13) | 1.02 (0.93–1.13) | 4.49 (3.74–5.39) | 2.81 (2.49–3.16) | 6.95 (5.73–8.43) |
| Behavioral syndromes (F50–F59) | 1.12 (1.00–1.25) | 5.56 (4.90–6.31) | 2.71 (2.26–3.26) | 7.18 (5.70–9.06) | 0.89 (0.80–1.00) | 4.22 (3.49–5.11) | 2.06 (1.71–2.49) | 8.32 (6.27–11.03) |
| Personality disorders (F60–F69) | 1.41 (1.09–1.82) | 6.31 (5.07–7.86) | 1.64 (1.37–1.96) | 5.88 (4.78–7.23) | 1.37 (0.99–1.90) | 6.42 (4.30–9.58) | 1.70 (1.34–2.16) | 7.55 (5.67–10.05) |
| Intellectual disability (F70–F79) | 7.38 (6.44–8.46) | 3.86 (1.98–7.50) | 4.00 (2.50–6.39) | 11.25 (5.70–22.22) | 11.79 (9.87–14.09) | 3.58 (1.28–9.99) | 4.53 (2.26–9.05) | NR |

| | Men | | | | Women | | | |
|---|---|---|---|---|---|---|---|---|
| | **Outside specialist psychiatric care** | | **Specialist psychiatric care** | | **Outside specialist psychiatric care** | | **Specialist psychiatric care** | |
| | **No SUD** | **SUD** | **No SUD** | **SUD** | **No SUD** | **SUD** | **No SUD** | **SUD** |
| Disorders of psychological development (F80–F89) | 1.58 (1.31–1.91) | 5.65 (4.07–7.84) | 1.40 (1.02–1.93) | 7.70 (5.21–11.37) | 2.28 (1.79–2.91) | 6.00 (3.31–10.89) | 2.22 (1.40–3.54) | 10.16 (5.01–20.63) |
| Behavioral and emotional disorders (F90–F98) | 1.27 (1.07–1.51) | 6.44 (5.24–7.90) | 1.51 (1.27–1.81) | 8.52 (7.02–10.34) | 1.23 (0.98–1.54) | 6.60 (4.58–9.51) | 2.02 (1.66–2.45) | 8.67 (6.36–11.82) |

Abbreviations: MRR, mortality rate ratio; NA, not applicable; NR, not reported; SUD, substance use disorder.

Diagnostic groups correspond to ICD-10 mental and behavioural disorder categories. Person-time was dynamically allocated according to time-varying psychiatric care setting and SUD status. MRRs were estimated separately for women and men using Poisson regression adjusted for age group, calendar period, region, and Charlson Comorbidity Index. For mental and behavioural disorders due to psychoactive substance use (F10-F19), estimates without SUD were not applicable. Diagnostic groups with insufficient deaths for stable estimation were not reported
